# Fluorosurfactants in groundwater increase the incidence of chronic health conditions among California Medicare beneficiaries

**DOI:** 10.1101/2024.02.25.24303330

**Authors:** Lucas M. Neas, William Steinhardt, K. Lloyd Hill, Riley Short, Elaine Hubal, Brian J Reich, Shu Yang, Alvin Sheng, Ana G. Rappold

## Abstract

**Background:** Per- and polyfluoroalkyl substances (PFAS) are persistent organic pollutants with emerging environmental and regulatory concerns.

**Objectives:** This study aimed to estimate the burden of PFAS exposures through ground water on the incidence of chronic health conditions among Medicare beneficiaries aged 65 years and older.

**Methods:** We estimated PFAS groundwater concentrations for every ZIP code tabulated area (ZCTA) in California counties where 25 percent or more of the population’s drinking water was derived from groundwater. We calculated the annual incidence of non-cancer chronic health conditions among 1,696,247 Medicare beneficiaries aged 65 and older by residential ZCTA over the seven-year study period (2011-2017). A Poisson regression model was used to estimate associations between PFAS groundwater concentration and chronic condition incidence with an offset for the number of beneficiary-years at risk and adjusting for bias due to non-random sampling of wells, use of groundwater for drinking water, demographic characteristics, and lung cancer incidence as a control for smoking.

**Results:** Results suggest an association between a 10 ng/L increment in PFAS contaminated groundwater and chronic health conditions including hypertension (+1.15%, 95% confidence interval (CI) 1.01, 1.30), chronic kidney disease (+0.83%, 95% CI 0.68, 0.99) and cataracts (+1.50%, 95% CI 1.35, 1.66).

**Discussion:** This small increment in the incidence rate would produce an additional 1,700 new cases of hypertension each year in the study population.

## Introduction

The rising concern over per- and poly-fluoroalkyl substances (PFAS) in drinking water in the United States has prompted the federal government to identify scientific challenges that must be addressed to understand and address human health impacts (National Science and Technology Council 2023). Potential adverse health effects from PFAS include impacts on fetal development, testicular and kidney cancer, liver tissue damage, suppressed immune function, and cholesterol levels changes (US Environmental Protection Agency 2021). While there is scientific consensus that there are significant health effects associated with perfluorooctanoic acid (PFOA) and perfluorooctane sulfonic acid (PFOS), the full scope of health effects in the population related to the larger set of PFAS measured in drinking water remains unclear.

Recent systematic evidence maps conducted by EPA researchers found limited epidemiological studies on cardiovascular endpoints related to PFAS exposure (Carlson, Angrish et al. 2022, Radke, Wright et al. 2022). Most epidemiological studies focused on acute clinical outcomes, such as heart attack-related hospitalizations, with few examining chronic conditions like hypertension, hyperlipidemia, or type-2 diabetes. These reviews identified few epidemiologic studies of cardiovascular endpoints in cross-sectional studies in the general population and even fewer in cohort studies. Most of these epidemiological studies were of acute clinical endpoints, such as deaths and hospitalizations for myocardial infarctions, and few have examined the effect of exposure on development of chronic health conditions such as the incidence of new diagnoses of major cardiovascular conditions, such as hypertension, hyperlipidemia, or type-2 diabetes.

Relative to the widespread use of these chemicals and known health effects, the evidence for population level health burden is still sparse, primarily due to the scarcity of exposure data. Although fluorosurfactants (principally PFOS and PFOA) have been detected in drinking water supplies throughout the US (Hu, Andrews et al. 2016, Guelfo and Adamson 2018), the assessment of the public health burden has been challenging; the exposures have not been systematically measured until recent history and PFAS is not generally measured in water used for consumption such as tap water. Simultaneously, PFAS exposures are both ubiquitous and differential with respect to many known determinants of chronic health conditions, such as income, race, sex, and occupation and there is an extensive lag time between exposure and the development of disease.

This study aims to assess the burden of chronic health conditions associated with PFAS exposure in groundwater well sources of drinking water. More specifically, the study estimates the risk associated with the annual incidence of common chronic health conditions among Medicare beneficiaries and identifies outcomes of highest concern for burden assessment at the population level. Propensity scores are employed to account for non-random sampling of wells and non-random use of groundwater for drinking. Additionally, several chronic health conditions are assessed as negative controls to identify potential unmeasured confounding related to smoking, frailty, and social isolation. The analysis quantifies outcomes with strongest association in response to PFAS exposure, the impact of each potential source of bias and offers recommendations for future analytical approaches.

## Methods

To estimate the burden of chronic health conditions associated with PFAS exposure we identified health outcomes from Centers for Medicare and Medicaid Services’ Chronic Conditions Warehouse, and exposure levels in groundwater well sources of drinking water. Additionally, areal and well level information about sociodemographic population characteristics, landcover types, and the presence of known and potential PFAS sources were used to control for confounding.

### Health Outcome Data

We obtained individual-level information on chronic health conditions among Medicare beneficiaries from the Centers for Medicare and Medicaid Services’ Chronic Conditions Warehouse. For each Medicare beneficiary, these annual files provide indicators for 27 chronic health conditions as well as their annually updated ZIP-code of residence (US Department of Health and Human Services 2023). Since Medicare is exceptional for those under 65 years, we restricted our analysis to Medicare beneficiaries aged 65 years and older for whom coverage is nearly universal.

The 27 chronic health conditions were separated into nine common conditions (cataract, chronic kidney disease, hypertension, congestive heart failure, hyperlipidemia, anemia, diabetes, rheumatoid/osteo arthritis, ischemic heart disease), eight less common conditions (chronic obstructive pulmonary disease (COPD), stroke, atrial fibrillations, glaucoma, hyperplasia, acquired hypothyroidism, osteoporosis, Alzheimer’s, and dementia), and three negative controls (lung cancer, depression, and hip fracture). We excluded from the analysis seven outcomes that averaged less than one incident case per year in a ZIP code: prostate cancer, breast cancer, endometrial cancer, colorectal cancer, asthma, Alzheimer’s, and acute myocardial infarction. We used indicators of each chronic health condition to calculate the annual incidence of newly diagnosed conditions over the seven-year period (2011-2017) for each ZIP code of residence.

For each ZIP code of residence, each unique beneficiary with an incident chronic health condition at any time during the seven-year study period contributed one count to the incidence of the corresponding condition. Person-years at risk were calculated in a similar manner; for each ZIP code of residence, each unique beneficiary contributed a beneficiary-year at risk for every year that they did not have the condition as well as the year in which they were first diagnosed.

Medicare beneficiary residential ZIP codes (postal delivery routes assigned by the US Postal Service) were mapped to both ZIP code population weighted centroids (US Department of Housing and Urban Development 2023) and ZIP Code Tabulated Areas (ZCTAs) (US Bureau of the Census 2023). In this process, multiple ZIP codes may have been mapped to the same ZCTA (2,444 ZIP codes were mapped to 1,756 ZCTAs).

### PFAS Groundwater Concentrations

PFAS concentrations in groundwater were obtained from California’s Groundwater Ambient Monitoring and Assessment (GAMA) program, a collaboration between the U.S. Geological Survey and California’s State Water Resources Control Board, which began testing domestic and public supply wells for PFAS in 2018 (US Geological Survey 2023). Well location data and PFAS sampling results were downloaded from the GAMA website for the period 2018 until 2021, however most samples were analyzed in recent periods (California Water Boards 2023).

Individual wells may have been tested several times for multiple PFAS species and all results were reported in nanograms per liter (ng/L). Values reported as less than the detection limit were interpreted as non-detects and converted to zero before calculating exposure; however, minimum detection limits varied by PFAS testing method. Maximum PFAS concentrations at individual wells were aggregated to the ZCTA-level as an inverse squared distance weighted concentration averaged across all wells within 20-km of the ZCTA population centroid, with wells closest to the centroid contributing the most weight.

### Sociodemographic Data

Population characteristics, including age, race, and income, were downloaded from the US Census Bureau’s 5-year American Community Survey (ACS-5) for all ZCTAs and census tracts in California for the year 2015 (US Bureau of the Census 2023). Additional sociodemographic data were retrieved from California’s Office of Environmental Health Hazard Assessment’s CalEnviroScreen 4.0 (CES) dataset which is an interactive mapping tool and database of environmental, socioeconomic, and public health data (California Office of Environmental Health Hazard Assessment 2023). The CES data were available by census tract, and for the purpose of the analysis they were averaged for all census tracts within each ZCTA as well as linked to the wells found within the census tract.

### PFAS Source

Locations of industries such as airports, fire training facilities, chemical manufacturing sites, and waste management sites registered through USEPA’s Enforcement and Compliance History Online (ECHO) database and included in EPA’s PFAS Analytics Tools were treated as potential PFAS sources in this study (‘PFAS Sources’, 23 variables, Table S1) (US Environmental Protection Agency 2023). Although, except for most military installations, this dataset does not include any actual data or confirmation about each facility’s PFAS use or discharge, these industry points are considered to have some relevance to PFAS use or discharge. Potential PFAS sources were counted within well site buffers (1- and 5-km) and ZCTAs by industry sector, and distances to the nearest PFAS industry source were measured from well sites. The number of PFAS industry sources were included as predictors of the exposure and were not considered to be on the pathway between PFAS concentration and health outcomes.

### Landcover Data

Land use and landcover data were obtained from the Multi-Resolution Land Characteristics Consortium’s 2016 National Land Cover Dataset (NLCD) (Dewitz and Survey 2021, Wickham, Stehman et al. 2021). The NLCD uses Landsat imagery to classify landcover into 16 categories at a 30m resolution for the entire US. The NLCD Land Cover Change Index raster dataset, which measures changes in landcover types between 2001-2019, was also included in this analysis. The NLCD also provides a raster of urban imperviousness that assigns each 30m pixel a value of percent developed impervious cover, for example, pavement and rooftops. Landcover data were summarized within California’s ZCTAs as well as within 1- and 5-km buffers around each well site. Local soil characteristics, including percent organic matter, percent clay, and pH, were summarized at each well site with data from the POLARIS database (Chaney, Wood et al. 2016, Moro Rosso, de Borja Reis et al. 2021). Elevation (meters above sea level) was estimated at each well site using the “elevatr” package in R (Hollister, Shah et al. 2022).

### Statistical approach

We used Poisson regression (“glm” package in R) (R Core Team 2022) to model the incidence of chronic health conditions as a function of PFAS concentrations (ng/L) in the groundwater. We first restricted study area to regions where at least 25 percent of drinking water was sourced from groundwater wells. This restriction was not based on PFAS groundwater concentrations nor incidence of chronic health conditions. However, to control for potential selection bias and for confounding by common causes introduced by choice of drinking water source we developed a propensity score (‘GW PS’). Second, we consider the possibility that wells were not tested at random and may have been differentially sampled with respect to unknown factors. To adjust for selection bias due to preferential sampling of wells, we also developed a well testing propensity score (‘Testing PS’) defining the probability of a well being tested based on measured covariates. Both propensity scores were included in the model as natural splines with three degrees of freedom.

Additionally, three population-level variables that are known determinants of health were added to the regression (‘Regional Determinants of Health’; percent of workforce employed in agriculture, percent Hispanic ethnicity, and percent with a disability). The prevalence of smoking (‘Smoking’) was unknown, both for the general population and for Medicare beneficiaries. Smoking is known to be the overwhelming cause of lung cancer and any contribution of PFAS must be orders of magnitude less, so we used lung cancer incidence as a surrogate to account for the unmeasured variation in the prevalence of smoking across the ZCTAs in the study area.

Finally, we adjusted for two sets of covariates that may predict the exposure: covariates related to the known and potential sources of PFAS (‘PFAS Sources’, 23 variables, Table S2) and covariates that were indicative of PFAS concentrations (‘PFAS Covariates’, 10 variables, Table S2).

We also considered depression and hip fracture, as negative controls to assess residual confounding by unknown or omitted sources of confounding. These outcomes have no known biological mechanisms that link the incidence of disease to PFAS exposure. However, we expect there may be other factors associated with negative controls and PFAS that are also likely to confound the association with other outcomes of interest. Therefore, the common causes of exposure to PFAS and outcome are hypothesized to be like the common causes of exposure to PFAS and incidence of chronic conditions. These factors may be directly or indirectly measured through variables describing the residential location, type of employment, proximity to sources, demographic characteristics, etc. Since social isolation is the major risk factor for depression, we believe that there exists no biological relationship between PFAS and depression. Any residual association between PFAS and depression would be evidence of residual confounding. We did not find literature reporting a direct link between hip fracture with PFAS exposure. The major risk factor for hip fracture is fall and age, but factors such as arthritis and osteoporosis may contribute to the severity of outcomes associated with hip fracture. Those outcomes are analyzed separately.

### Propensity score for PFAS testing

Groundwater wells were not randomly selected for PFAS testing which can potentially lead to selection bias in the analysis. To account for potential selection bias, we estimate the probability of each well being selected for PFAS testing (testing propensity score). We take advantage of the large and comprehensive characterization of residential and potential PFAS sources within 1-km and 5-km radii of each well and use extreme gradient boosting, XGBoost algorithm (Chen and Guestrin 2016), to predict the probability of a well being tested for PFAS. More specifically, the following variables were summarized at the well (buffer) level and used in the XGBoost algorithm: the Californian hydrologic region (https://gis.data.ca.gov/datasets/2a572a181e094020bdaeb5203162de15/explore) that the well is in, land use and landcover variables from the NLCD, location of potential industrial PFAS sources (EPA PFAS Analytics Tool), population characteristics (CES and US Census, ACS), soil characteristics (POLARIS), and elevation.

The XGBoost model was fit in two stages using the test-set validation approach. The model in the first stage was fit on a random subset containing 90% of wells, with the optimal number of gradient boosting rounds determined via early stopping with ten rounds, using the other 10% of wells as a validation set. That is, the optimal round is the last round after which performance on the validation set doesn’t improve for ten rounds. The model in the second stage was then fit on all wells, with the optimal number of rounds found in the first stage. The other parameters of the model are set to the defaults as in the “xgboost” package in R (https://xgboost.readthedocs.io/en/stable/parameter.html). This model was then used to predict propensity scores for all the wells. We aggregated propensities to the ZCTA-level as an inverse distance weighted concentration averaged across all PFAS-tested wells within 20-km of the ZCTA population centroid, as with PFAS concentrations.

### Propensity score for drinking water from ground sources

We consider groundwater consumption in the epidemiological model for PFAS in groundwater. While we estimated 22% of Californians rely on groundwater as their primary source of drinking water, this proportion varies considerably by county. In the study area, the overall proportion was 54%. Data on public water systems, including population served and whether sourced from groundwater or surface water, were downloaded from California’s Public Water System Information Dataset (PWSID) (California State Water Resources Control Board. 2019). These water-source estimates were combined with estimates of population served by private drinking water wells (Johnson, Belitz et al. 2019) to calculate percent of county population drinking from groundwater. We restricted the study region to ZCTAs with population centroids within the 40 counties where 25% or more of the population rely on groundwater for their drinking water.

To account for potential selection bias from restricting our analysis to the specified ZCTAs, we calculate the second propensity score for each ZCTA (probability of selection in the study region). We use extreme gradient boosting (Chen and Guestrin 2016) to calculate the propensity scores, as earlier. However, for the two-stage test-set validation approach, the model was trained on 80% of the ZCTAs and validated on the other 20%; the validation set has a larger proportion to adjust for the smaller number of ZCTAs. We predicted these propensity scores conditional on ZCTA-level variables: land use and landcover variables (NLCD), location of potential industrial PFAS sources (EPA PFAS Analytics Tool), and population characteristics (CES and US Census, ACS).

### Statistical models

For each outcome of interest, we fit seven models (M1-M7) for weighted PFAS concentrations as a treatment variable of interest. M1 (Base Model) used the log count of the total Medicare beneficiaries (population-at-risk) as an offset. The subsequent models were cumulative adjustments to the basic model: M2 (+ Testing PS) adds the PFAS-well testing propensity score to control for potential selection bias; M3 (+ GW PS) adds the propensity score for the proportion of drinking water from groundwater; M4 (+ Regional Determinants of Health) adds three major regional health determinants for the total population, including percent working in agriculture industry, percent Hispanic and percent with disability; M5 (+ Smoking) adds lung cancer incidence as a surrogate for smoking prevalence. Finally, model M6 (+ PFAS Sources) adds known industrial PFAS sources (Supplemental Table S1) and M7 (+ PFAS Covariates) adds additional covariates (not including sources) that were related to high levels of PFAS (Supplemental Table S2).

Sensitivity analysis was conducted with respect to the exposure metric. In addition to the weighted metric used in the main analysis, we also considered unweighted, and propensity weighted metric. The propensity weighted metric was constructed as 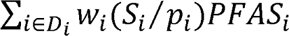, where *w_i_* is the distance weight of the well to the population centroid of ZCTA, *S_i_* is the indicator of a sampled location, and *p_i_* is the propensity score of the sampling and the summation is over sampled locations in *D_S_*. Rescaling the original propensity score 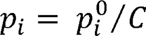, where 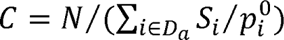 provides a property of 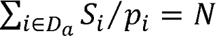. As a result, the constructed propensity weighted metric approximates the weighted average of PFAS had all the wells in the ZCTA *D_S_* been sampled 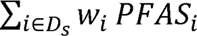. This metric estimates the total exposure for this ZCTA and adjusts for preferential sampling by weighting.

Results are reported as the percent difference in the annual incidence of a chronic condition for a 10.0 ng/L increment in estimated PFAS groundwater concentrations, which corresponds to the 80^th^ percentile of the PFAS distribution in the study area. To recapitulate, PFAS groundwater concentrations are estimated for each ZCTA as the inverse distance-squared weighted mean of the well-specific maximum PFAS-species concentration for tested wells within 20-km of the ZTCA population centroid.

## Results

The study cohort included 1.7 million Medicare beneficiaries aged 65 years and older living within 523 ZCTAs (Figure 1). The greatest reduction in the study cohort was the restriction of the study to beneficiaries residing in California counties with over 25% of their drinking water derived from groundwater, which excluded many of the major urban centers (Figure 1 inset map). Compared to all of California, the population of the study area had greater proportions of Hispanic ancestry, with disabilities and employed in agriculture (Table 1). The study area had a slightly higher incidence of the eight most prevalent chronic health conditions, but a similar incidence of lung cancer, which was used as a proxy for smoking prevalence (Table 1).

**Figure 1.**
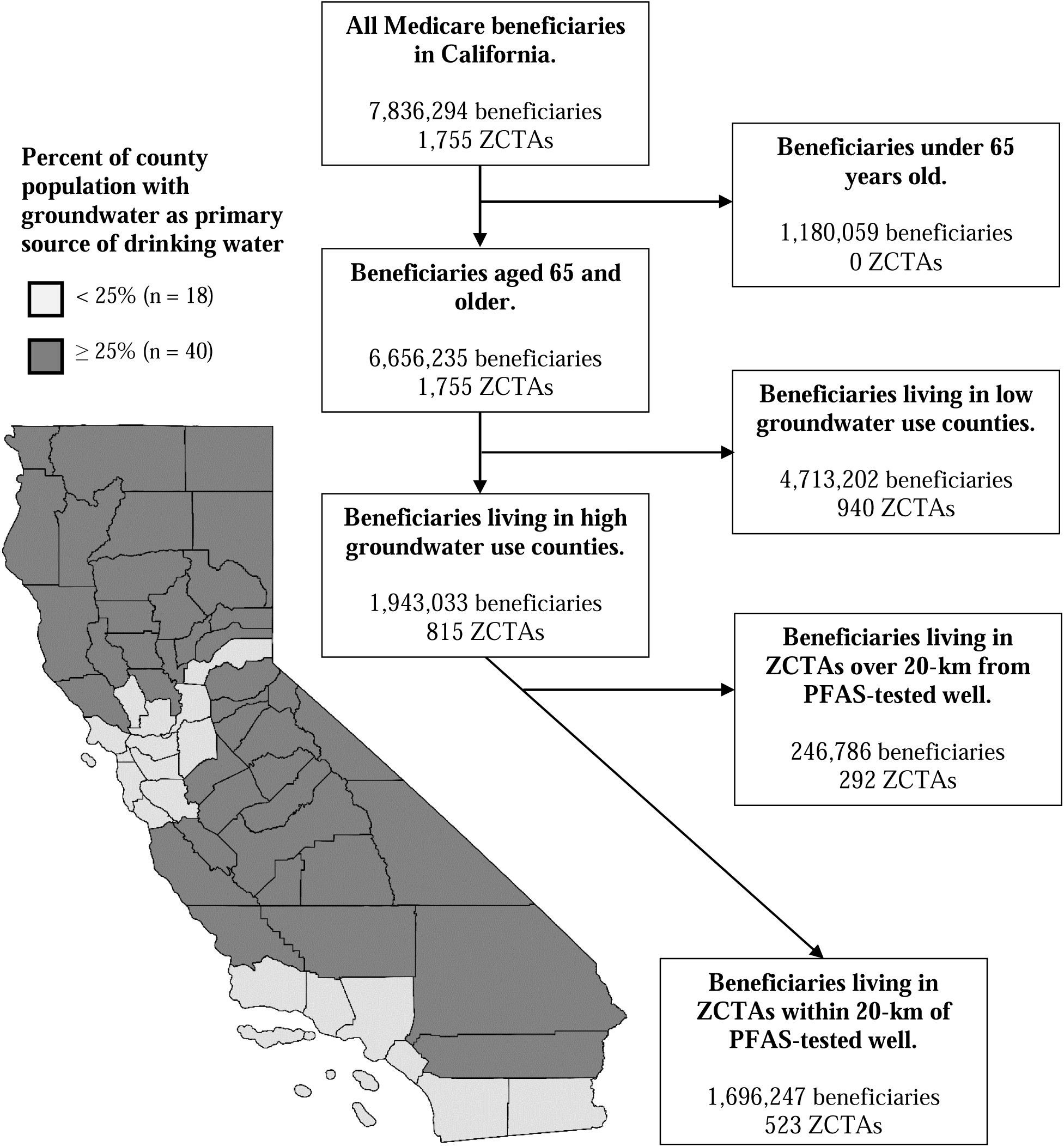
Flow diagram showing restrictions to cohort in number of Medicare beneficiaries and ZIP Code Tabulation Areas (ZCTAs) with inset map of California counties showing proportion of population supplied with drinking water derived from groundwater: 2011-2017.

**Table 1.**
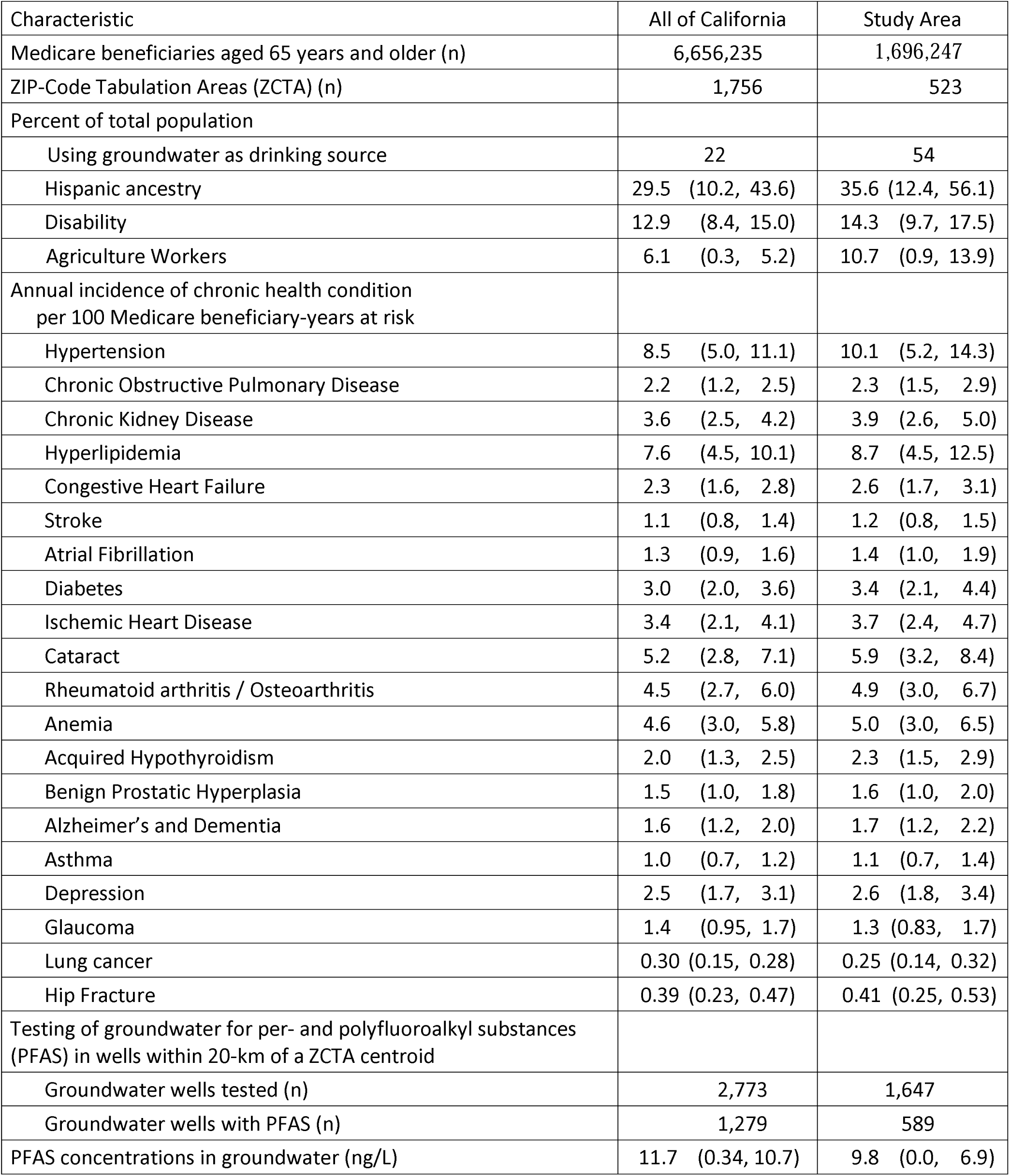
Characteristics of study area in comparison with all of California: 2011-2017. The study area was restricted to California counties where 25% or more of the population’s drinking water was supplied from groundwater. Unless otherwise specified, results are presented as means and interquartile ranges across Zip Code Tabulation Areas (ZCTA).

| Characteristic | All of California | Study Area |
| --- | --- | --- |
| Medicare beneficiaries aged 65 years and older (n) | 6,656,235 | 1,696,247 |
| ZIP-Code Tabulation Areas (ZCTA) (n) | 1,756 | 523 |
| Percent of total population |  |  |
| Using groundwater as drinking source | 22 | 54 |
| Hispanic ancestry | 29.5 (10.2, 43.6) | 35.6 (12.4, 56.1) |
| Disability | 12.9 (8.4, 15.0) | 14.3 (9.7, 17.5) |
| Agriculture Workers | 6.1 (0.3, 5.2) | 10.7 (0.9, 13.9) |
| Annual incidence of chronic health condition<br>per 100 Medicare beneficiary-years at risk |  |  |
| Hypertension | 8.5 (5.0, 11.1) | 10.1 (5.2, 14.3) |
| Chronic Obstructive Pulmonary Disease | 2.2 (1.2, 2.5) | 2.3 (1.5, 2.9) |
| Chronic Kidney Disease | 3.6 (2.5, 4.2) | 3.9 (2.6, 5.0) |
| Hyperlipidemia | 7.6 (4.5, 10.1) | 8.7 (4.5, 12.5) |
| Congestive Heart Failure | 2.3 (1.6, 2.8) | 2.6 (1.7, 3.1) |
| Stroke | 1.1 (0.8, 1.4) | 1.2 (0.8, 1.5) |
| Atrial Fibrillation | 1.3 (0.9, 1.6) | 1.4 (1.0, 1.9) |
| Diabetes | 3.0 (2.0, 3.6) | 3.4 (2.1, 4.4) |
| Ischemic Heart Disease | 3.4 (2.1, 4.1) | 3.7 (2.4, 4.7) |
| Cataract | 5.2 (2.8, 7.1) | 5.9 (3.2, 8.4) |
| Rheumatoid arthritis / Osteoarthritis | 4.5 (2.7, 6.0) | 4.9 (3.0, 6.7) |
| Anemia | 4.6 (3.0, 5.8) | 5.0 (3.0, 6.5) |
| Acquired Hypothyroidism | 2.0 (1.3, 2.5) | 2.3 (1.5, 2.9) |
| Benign Prostatic Hyperplasia | 1.5 (1.0, 1.8) | 1.6 (1.0, 2.0) |
| Alzheimer's and Dementia | 1.6 (1.2, 2.0) | 1.7 (1.2, 2.2) |
| Asthma | 1.0 (0.7, 1.2) | 1.1 (0.7, 1.4) |
| Depression | 2.5 (1.7, 3.1) | 2.6 (1.8, 3.4) |
| Glaucoma | 1.4 (0.95, 1.7) | 1.3 (0.83, 1.7) |
| Lung cancer | 0.30 (0.15, 0.28) | 0.25 (0.14, 0.32) |
| Hip Fracture | 0.39 (0.23, 0.47) | 0.41 (0.25, 0.53) |
| Testing of groundwater for per- and polyfluoroalkyl substances<br>(PFAS) in wells within 20-km of a ZCTA centroid |  |  |
| Groundwater wells tested (n) | 2,773 | 1,647 |
| Groundwater wells with PFAS (n) | 1,279 | 589 |
| PFAS concentrations in groundwater (ng/L) | 11.7 (0.34, 10.7) | 9.8 (0.0, 6.9) |

Among the more than 20,000 wells extracted from the GAMA database, only 2,773 (14%) were tested for PFAS. Statewide, the highest PFAS groundwater concentrations were observed at wells near the most urbanized areas with low groundwater usage for drinking which were not located within the study area (for example, Los Angeles, San Francisco, and San Diego) suggesting that regions less reliant on ground sources of drinking water have more contamination and are tested more often. Within the study area, 1,647 wells were tested for PFAS and 589 (36%) had detectable levels of PFAS, with higher PFAS concentrations observed at wells near or within urban areas (for example, Fresno, Redding, and San Luis Obispo). Although the highest PFAS concentrations were outside the study area, the overall distributions of PFAS concentrations within the study area were remarkably similar to those of California overall (Table 1).

The exposure metric is calculated using the maximum sample at each well regardless of PFAS species. As such, only a handful of the tested PFAS species were represented as the exposure metrics, since 98% of wells’ maximum PFAS measurements were from just five species – PFOS, PFHXSA, PFOA, PFHA, and PFBSA (Table S4). These five species are either perfluoroalkyl sulfonic or perfluoroalkyl carboxylic acids with chain lengths ranging from 4 to 8 carbons. Four additional PFAS species (PFBTA, PFPES, PFUNDCA, NMEFOSAA) were also detected in study area wells, but were never the maximum measured species for any well.

Based on our analysis of testing propensity, the average probability of a groundwater well being tested for PFAS was 89.1% for wells with detectable levels of PFAS and 83.2% for wells without detectable levels of PFAS, indicating that contaminated wells were more likely to be tested than non-contaminated wells. Variables most influential in prediction of well testing were increased land development (“developed, medium-intensity”, an NLCD category) within 5-km of the well, presence of 1,2,3-trichloropropane (an industrial solvent), decreased levels of chromium 6 and uranium, and shorter distance to national defense industry sites. The presence of PFAS sources explained only 3% of variability (R^2^) in PFAS concentration across wells.

The incidences of chronic health conditions were associated with a 10 ng/L increment in PFAS groundwater concentrations with the strongest associations observed for hypertension 1.15% (1.01, 1.30), cataract 1.50% (1.35, 1.66), chronic kidney disease 0.83% (0.68, 0.99), and COPD 1.21% (1.01, 1.42) for the final covariate model (Table 2, Table S4). Adjustments for PFAS testing propensity and for varying use of groundwater for drinking water produced little change in estimates, but the three regional health covariates and smoking (that is, the ZCTA-specific incidence of lung cancer) were strong confounders as shown by the change in estimates. Ischemic heart disease as a chronic health condition represents a cohort of heart attack survivors and these results suggest that PFAS may have a negative impact on heart attack survival. Adjustments for PFAS sources (23 degrees of freedom) and for other covariates (10 degrees of freedom) showed little evidence of additional confounding although addition of PFAS sources generally increased the effect estimates.

**Table 2.** Estimated percent difference (and 95% confidence intervals) in annual incidence of selected chronic health conditions for a 10 ng/L increment in distance weighted mean of per- and polyfluoroalkyl substances (PFAS) groundwater concentration by epidemiologic model: 1,696,247 Medicare beneficiaries over age 65 living in California counties with more than 25% of drinking water from groundwater, 2011-2017.

|  | Epidemiologic model |  |  |  |  |  |  |
| --- | --- | --- | --- | --- | --- | --- | --- |
|  | Base Model | + Testing PS | + GW PS | + Regional Determinants of Health | + Smoking | + PFAS Sources | + PFAS Covariates "Final Model" |
| Model Degrees of Freedom | 2 | 5 | 8 | 11 | 14 | 37 | 47 |
| Hypertension | 0.92<br>(0.78, 1.07) | 0.66<br>(0.51, 0.81) | 0.78<br>(0.63, 0.93) | 2.00<br>(1.86, 2.14) | 1.12<br>(0.99, 1.26) | 1.25<br>(1.11, 1.39) | 1.15<br>(1.01, 1.30) |
| Chronic Obstructive Pulmonary Disease | 1.40<br>(1.2, 1.6) | 1.32<br>(1.12, 1.53) | 1.69<br>(1.48, 1.9) | 1.87<br>(1.67, 2.07) | 1.04<br>(0.85, 1.24) | 1.32<br>(1.12, 1.53) | 1.21<br>(1.01, 1.42) |
| Chronic Kidney Disease | 0.66<br>(0.51, 0.82) | 0.59<br>(0.44, 0.75) | 0.77<br>(0.62, 0.93) | 1.32<br>(1.17, 1.47) | 0.73<br>(0.58, 0.88) | 0.84<br>(0.68, 0.99) | 0.83<br>(0.68, 0.99) |
| Cataract | 2.37<br>(2.21, 2.53) | 2.19<br>(2.02, 2.35) | 1.97<br>(1.80, 2.14) | 2.52<br>(2.37, 2.68) | 1.68<br>(1.53, 1.83) | 1.73<br>(1.57, 1.88) | 1.50<br>(1.35, 1.66) |
| Depression | 0.43<br>(0.24, 0.62) | 0.36<br>(0.16, 0.55) | 0.34<br>(0.14, 0.54) | 0.58<br>(0.39, 0.77) | 0.15<br>(-0.04, 0.33) | 0.20<br>(0.00, 0.39) | 0.09<br>(-0.10, 0.28) |
| Hip Fracture | 1.01<br>(0.59, 1.43) | 1.00<br>(0.57, 1.43) | 0.99<br>(0.56, 1.43) | 0.78<br>(0.36, 1.19) | 0.26<br>(-0.15, 0.67) | 0.34<br>(-0.08, 0.77) | 0.11<br>(-0.32, 0.54) |

Base model: A conditional Poisson model for incidence of chronic health condition in a Zip Code Tabulation Area (ZCTA) and distance-weighted mean of well-specific maximum PFAS concentration within 20-km of ZCTA population centroid with an offset for beneficiary-years at risk.
+ Testing PS: Previous model with a propensity score (PS) for PFAS testing based on land-use near well.
+ GW PS: Previous model with a propensity score (PS) for county indicators of use of groundwater (GW) for drinking water.
+ Regional Determinants of Health: Previous model with percent working in agriculture, percent Hispanic, and percent with disability.
+ Smoking: Previous model plus the incidence of lung cancer as an estimate for smoking prevalence.
+ PFAS Sources: Previous model plus 23 industrial sector-specific counts of PFAS sources in ZCTA (supplemental table S1).
+ PFAS Covariates: Previous model plus 10 covariates indicative of high PFAS concentrations (supplemental table S2).

Associations for other non-cancer chronic health conditions generally had small but positive estimated risk, except for asthma and ischemic heart disease (Figure 2). The associations for some outcomes were sensitive to one largest PFAS value observed in Mariposa County. After removing this maximum value, associations between PFAS and anemia, asthma, and diabetes were null. For all other outcomes estimates of risk were consistent and similar in magnitude (Figure S3).

**Figure 2.**
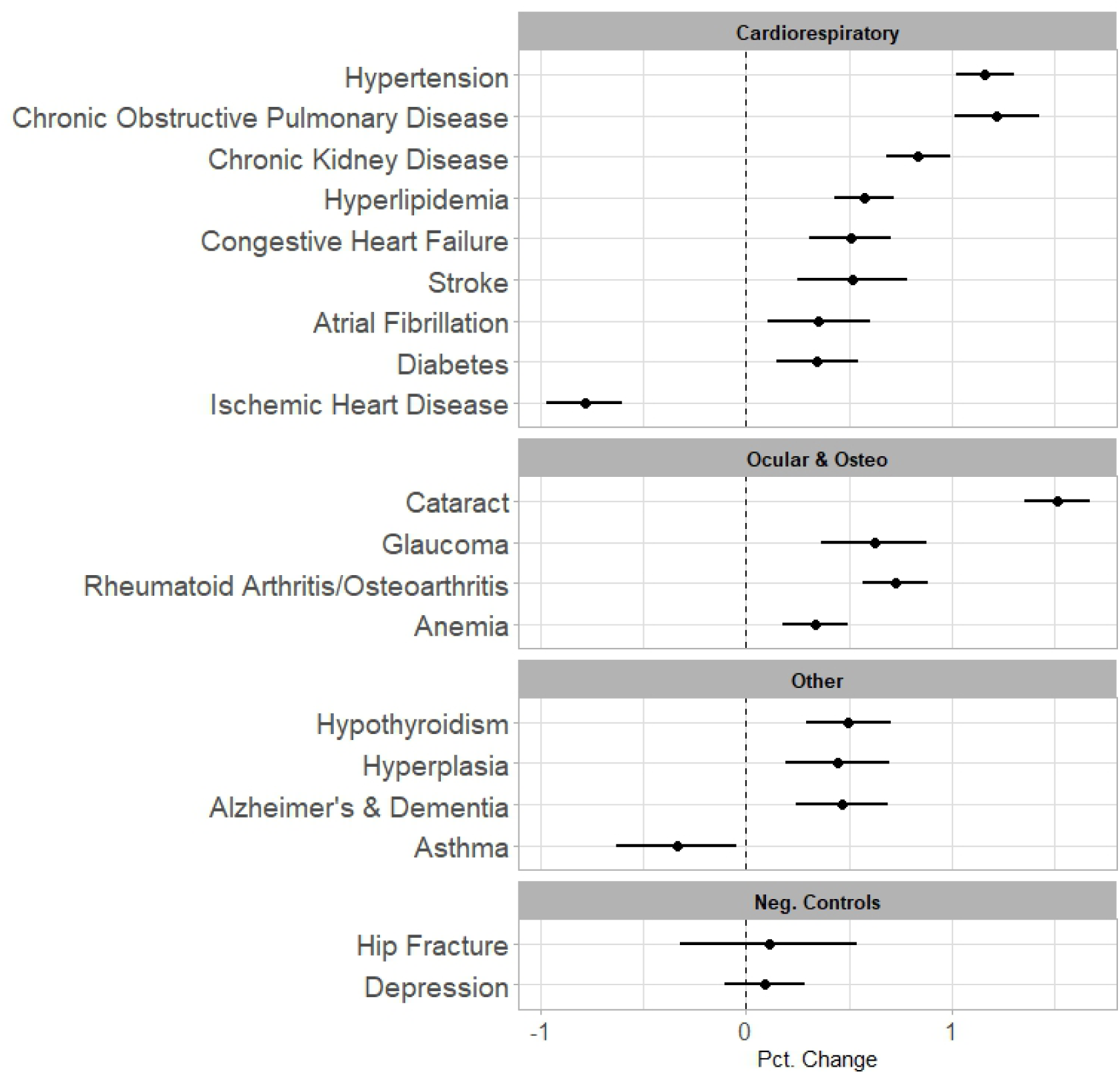
Estimated percent difference in incidence of 19 chronic health conditions for a 10 ng/L increment in distance-weighted mean of per- and polyfluoroalkyl substances (PFAS) groundwater concentration (and 95% confidence intervals) for final epidemiologic model: 1,696,247 Medicare beneficiaries over age 65 living in California counties with more than 25% of drinking water from groundwater, 2011-2017.

Many of these chronic health conditions share common etiologies and these results may reflect shared pathways of action. We performed a mediation analysis to assess whether PFAS associations were mediated by the strong association between PFAS and hypertension, a known risk factor for other chronic health conditions. Hypertension is also the most prevalent condition and has the largest rate of co-occurrence with other conditions. With the addition of hypertension to the model, the PFAS direct effect was reduced to 0.56 (95% CI 0.40, 0.71) for chronic kidney disease, 0.76 (95% CI 0.61, 0.92) for cataract, and 0.94 (95% CI 0.73, 1.15) for chronic obstructive pulmonary disease. The proportion of the total effect mediated by hypertension was greatest for cataracts (49%), chronic kidney disease (33%), and chronic obstructive pulmonary disease (22%). These results suggest that hypertension mediated a considerable portion, but not all, of the association between PFAS and these other chronic health conditions.

## Discussion

Exposure to PFAS is a national public health concern with a growing need for population level risk assessment. In this study, we characterized the association between PFAS in the groundwater wells and the incidence cases of chronic disease in a large population of Medicare beneficiaries aged 65 years and older residing in California between 2011 and 2017. The study area was a region where approximately 54% of population receives drinking water from ground water and the exposure was determined as a distance-weighted mean of the maximum PFAS concentrations at each groundwater well in the postal code of Medicare beneficiaries. We observed the strongest association for the incidence of hypertension, chronic kidney disease, COPD, and cataract; however, the incidence of other chronic diseases (congestive heart failure, stroke, atrial fibrillations, diabetes, glaucoma, osteoarthritis, anemia, acquired hypothyroidism, benign prostatic hyperplasia, and Alzheimer’s/dementia) were also positively associated with PFAS exposure.

Incidence of hypertension was increased by one percent (1.15, 95% CI 1.01, 1.30) for a 10 ng/L increment of PFAS. To place this small rate increment in context, the average annual incidence of hypertension in the study area was 10.1 percent or about 170,000 new cases of hypertension annually. A one percent increase in this rate would add 1,700 additional cases of hypertension each year among the approximately 1.7 million Medicare beneficiaries in the study area.

An association between PFAS and hypertension has been previously reported. Cardiovascular disease has been linked to PFAS by experimental and epidemiologic studies (Meneguzzi, Fava et al. 2021) (Meneguzzi, Fava et al. 2021, Wen, Wang et al. 2022). Hypertension and dyslipidemia have been linked to serum PFAS among Norwegian adolescents (Averina, Brox et al. 2021), US women (Ding, Karvonen-Gutierrez et al. 2022) and adolescents (Ma, Xu et al. 2019), and to PFAS drinking water contamination among North Carolina adults (Ward-Caviness, Moyer et al. 2022). Preeclampsia, acute onset hypertension in pregnant women, and other reproductive related outcomes were found to be associated with PFAS in two large systematic reviews (Gao, Ni et al. 2021, Rickard, Rizvi et al. 2022). This study is the first to provide an estimate of the burden of PFAS in groundwater on clinically reported hypertension among a general population aged 65 years and older. An adverse association between PFOS and preeclampsia (acute hypertension in pregnancy) was first reported for women enrolled in a study of a major industrial release of PFAS in the Mid-Ohio River Valley in the United States (Stein, Savitz et al. 2009, Avanasi, Shin et al. 2016). Adverse associations between PFAS and preeclampsia were also found in Sweden (Rylander, Lindh et al. 2020), Denmark (Birukov, Andersen et al. 2021), Canada (Borghese, Walker et al. 2020), United States (Preston, Hivert et al. 2022) and China (Tian, Zhou et al. 2023) but results from a Norwegian cohort were equivocal (Starling, Engel et al. 2014).

The results reported here are consistent with a previous study in North Carolina based on electronic health records (Ward-Caviness, Moyer et al. 2022). That study also reported similar associations between presence of any PFAS detected in the drinking water system with hypertension (OR 1.32 (1.15, 1.52)) and chronic kidney disease (OR 1.14 (0.87, 1.51)). In that study, the associations between PFAS and heart failure and diabetes were problematic due to smaller sample size, and cataract and COPD were not considered. Even though both studies were population based, they used different exposure metrics: a binary indicator of any PFAS detected in a water system versus more extensive PFAS testing of groundwater well concentrations.

Two outcomes had negative association with PFAS, asthma and ischemic heart disease. Both outcomes have previously been associated with PFAS but not necessarily in this age group. Here because we identify the outcomes as the incident cases that have occurred for the first time after age 65, we suspect that we may have conditioned on the population that is less likely to be newly diagnosed with these two outcomes, like the healthy survivor bias. More specifically, new incidence of asthma is rare in older adults while ischemic heart disease is diagnosed only in the surviving cases (∼75%).

The mediation analysis suggested that hypertension was an important mediator of the PFAS association with other chronic health conditions, especially for cataracts and chronic kidney disease but less so for chronic obstructive pulmonary disease. Even after consideration of hypertension, PFAS exposure still showed direct effects on these other conditions suggesting that the PFAS association is not entirely mediated by hypertension. The relationship of hypertension with cataracts and chronic kidney disease has previously been established (Yu, Lyu et al. 2014, Hamrahian and Falkner 2017). The causal relationship between hypertension and chronic obstructive pulmonary disease is more complex as the loss of lung structure may cause hypertension (Kim, Park et al. 2017). Nevertheless, the results of this study indicate that PFAS has a direct association with these chronic health conditions beyond that mediated by hypertension. Ward-Caviness et al. (Ward-Caviness, Moyer et al. 2022) similarly indicated that the health effects of the exposure to PFAS are not specific and may be associated with multiple chronic conditions.

### Strengths and weaknesses

This study has several strengths and weaknesses. For those over the age 65, Medicare coverage is essentially universal. Thus, data collected on new incidence of chronic disease is representative of the population in the same age group in the study region. A unique strength of Medicare beneficiary summaries is that the chronic diseases are tracked over all beneficiaries and the incidence and prevalence rates can be fully enumerated. Nevertheless, to our knowledge there have not been other studies of PFAS effects using beneficiary files.

In this analysis we payed close attention to various sources of potential bias including preferential sampling of PFAS and confounding. We carefully considered selection bias from the non-random sampling of wells for PFAS testing, but the adjustment by a testing propensity score showed minimal impact on the PFAS effect estimates. The restriction of the study area to California counties where more than 25% of drinking water comes from groundwater was also adjusted by a propensity score for groundwater use and several demographic characteristics associated with the selection. The adjustment had minimal impact on the PFAS effect estimates (Table 2), but the reported findings ought not to be extrapolated to areas that were excluded from the analysis. For example, the region around Los Angeles has a number of wells with detected PFAS levels however, the region is estimated to have low reliance on the use of ground water sources for drinking. Additionally, some of the outcomes that had the largest risk estimates, such as cataract, have previously been noted to have both genetic and age dependent risk factors. Although conceivable that an association exists between average age in population and average PFAS concentration and that the outcome of interest is a surrogate for age, the risk estimates did not change after adjusting for age.

PFAS sources had a small suppression effect (MacKinnon, Krull et al. 2000), which when added to the model increased the effect estimates across most outcomes. The small explanatory power of PFAS sources suggest that presence of sources cannot act as an Instrumental variable and therefore cannot be a substitute for the direct exposure measurements. However, the addition of these variables can increase the magnitude and improve the overall model.

This study is a cross sectional analysis of chronic condition incidence and PFAS exposure in ground water used for drinking water, and as such is subject to several sources of potential confounding. The major confounders in the analysis were the three regional determinants of health (Hispanic ethnicity, disabilities, and agricultural employment) and the incidence of lung cancer as a surrogate for tobacco smoking (Table 2). Adjustment for the three regional variables increased the PFAS associations, while the adjustment for lung cancer reduced these associations. Among other limitations, smoking status of beneficiaries was not available and lung cancer incidence is used to characterize population level rates of smoking. Tobacco smoking does have a strong independent association with hypertension and partial adjustment for smoking using lung cancer incidence shows a large change in estimate, reflecting considerable confounding. The adjustment by the propensity scores for groundwater use showed less evidence of confounding. The widths of the confidence intervals for each chronic condition do not change much across the epidemiologic models (Table 2), providing no evidence of overfitting. This lack of variance inflation persists even for meaningful changes in the point estimates.

This study is an ecological study of a shared environmental exposure within the ZIP code of residence. Population level assessments are challenging due the lack of a comprehensive sampling strategy for PFAS. In this study, we used measurements for PFAS in groundwater from GAMA data repository of various sampling campaigns. While possible, measurement errors are unlikely to have varied across ZCTAs differentially with the incidence of chronic health conditions. Similarly, the clinical diagnosis of chronic health conditions supplied by the Medicare data is unlikely to have been influenced by PFAS groundwater concentrations. Finally, temporal misalignment between sampling of PFAS, exposure, and measured outcomes in these early stages of population level investigation is a knowledge gap and should not be overlooked.

Future research will benefit from enhanced measurements of PFAS in drinking water sources that is ongoing in many States. In present work we use the best available data on groundwater concentrations, but measures of drinking water exposure are likely to provide direct linkage. We found that adjustment for area-level lung cancer incidence was an important confounder, which implies that smoking is likely to be an important factor for consideration in future studies. The second most influential confounders included areal-level determinants of population health, which in the study area were % hispanic population, % disability, and % employed in agriculture. Adjustment with propensity scores for the non-random sampling of wells for PFAS testing and for differences in groundwater use for drinking water produced modest changes in the estimates but are important to consider in studies of ground water exposure. The impact of groundwater use was minimized by our restriction to California counties with at least 25% of drinking water derived from groundwater. Finally, the range of chronic conditions associated with PFAS indicated non-specific modes of action. Separation of PFAS associations into direct and indirect effects would be useful in resolving these pathways, as shown by our analysis using hypertension as a mediator.

## Conclusion

The findings of associations between PFAS in the groundwater and the incidence of hypertension, chronic obstructive pulmonary disease, chronic kidney disease, and cataracts among Medicare beneficiaries aged 65 years and older are consistent with prior clinical and epidemiologic research and are unlikely to be due to confounding, selection bias, or measurement errors. While a small percentage increase in annual incidence of clinical hypertension may seem like a small proportion, the impact of hypertension on subsequent morbidity and mortality suggests important public health consequences.

## Data Availability

All exposure data and analytical code will be posted after the manuscript is accepted. Health data is considered sensitive and cannot be posted publicly.

## Acknowledgments

Although this work has been reviewed for publication by the U.S. Environmental Protection Agency, it does not necessarily reflect the views and policies of the Agency. Reference herein to any specific commercial products, process, or service by trade name, trademark, manufacturer, or otherwise, does not necessarily constitute or imply its endorsement, recommendation, or favoring by the United States Government.

## Abbreviation

ZIP-code: Zone Improvement Plan Code
ZCTA: Zip Code Tabulation Area
EPA: Environmental Protection Agency
GAMA: Groundwater Ambient Monitoring and Assessment
ACS-5: American Community Survey
CES: CalEnviroScreen
ECHO: Enforcement and Compliance History Online
NLCD: National Land Cover Dataset
POLARIS: Probabilistic Remapping of Soil Survey Geographic Database
PWSID: Public Water System Information Dataset
COPD: Chronic Obstructive Pulmonary Disease
GW: Groundwater
PS: Propensity Score
glm: Generalized Linear Model
XGBoost: Extreme Gradient Boosting
IQR: Inner quartile range
ng/L: nanograms per liter
km: kilometer
m: meter
pH: Potential of Hydrogen
DAG: Directed Acyclic Graph
PFAS: Per- and polyfluoroalkyl sulfonic acids
PFOA: perfluorooctnoic acid
PFOS: perfluoroocane sulfonic acid
PFHXSA: perfluorooctanoic sulfonate
PFHA: perfluorohexanoic acid
PFBSA: perfluorobutanesulfonic
PFBTA: Perfluorobutanoic acid
PFPES: Perfluoropentanesulfonoic acid
PFUNDCA: Perfluoroundecanoic acid
NMEFOSAA: N-Methyl perfluorooctane sulfonamidoacetic acid

## Supplemental Figures and Tables

**Table S1.**
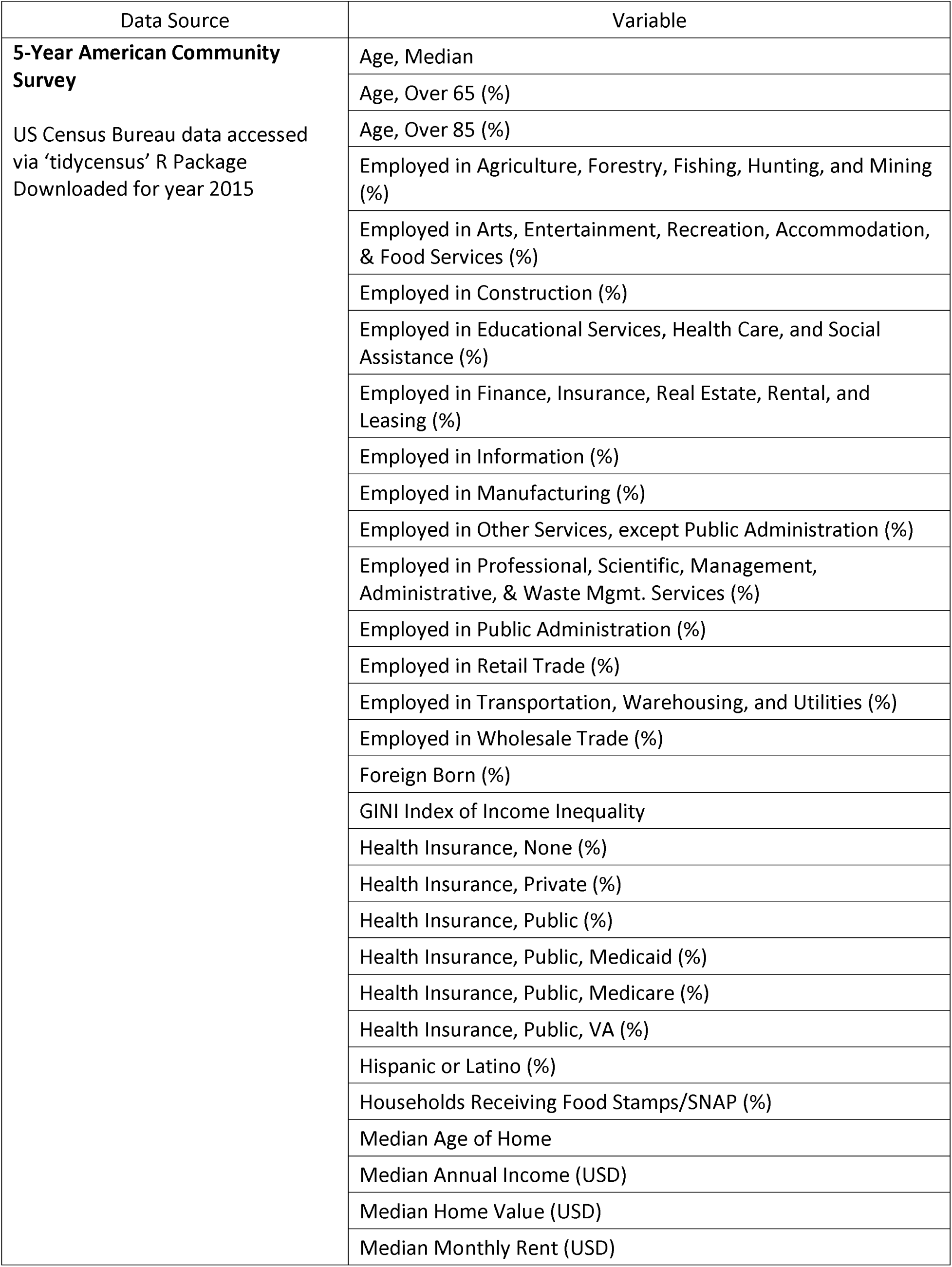
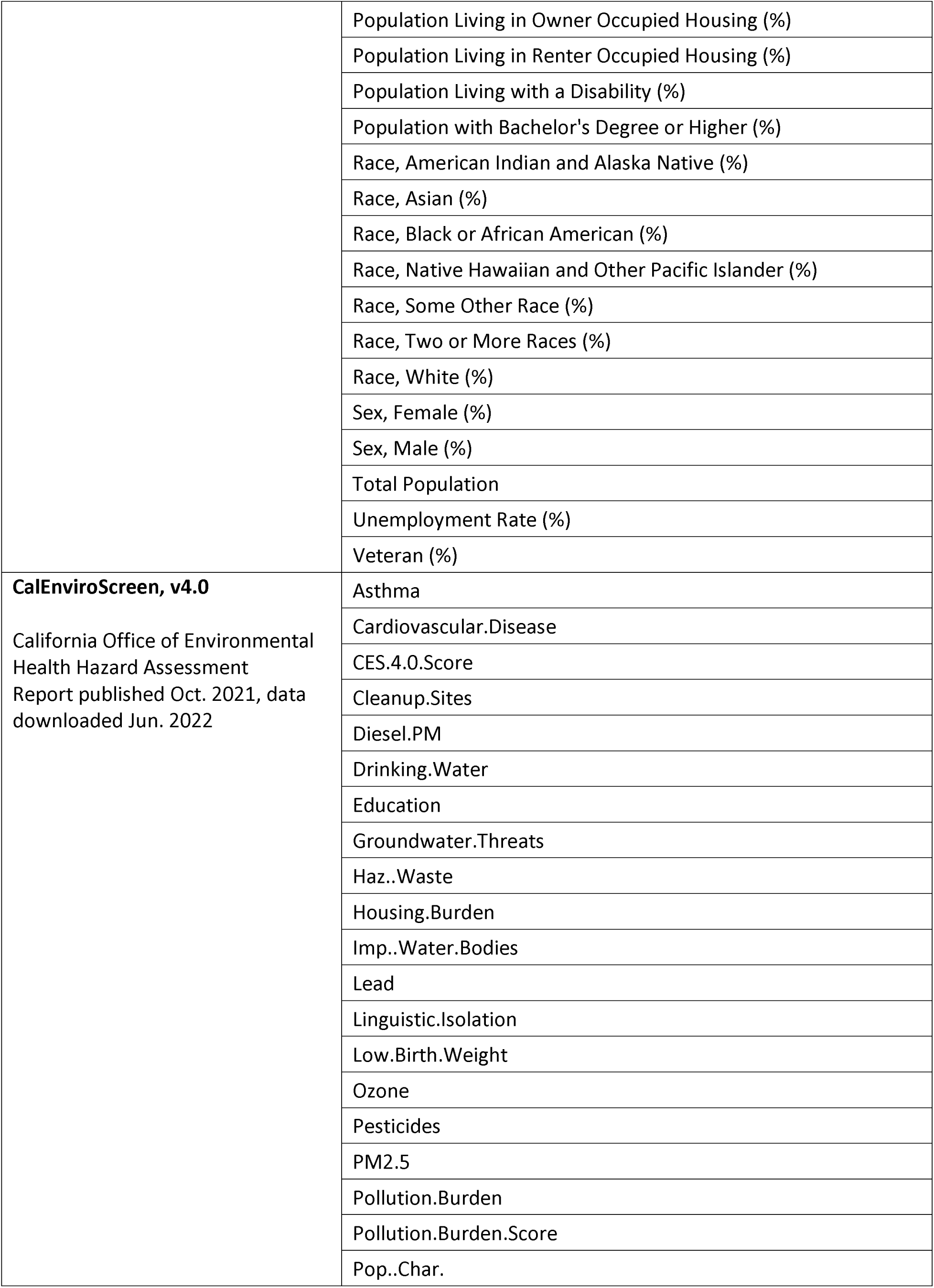
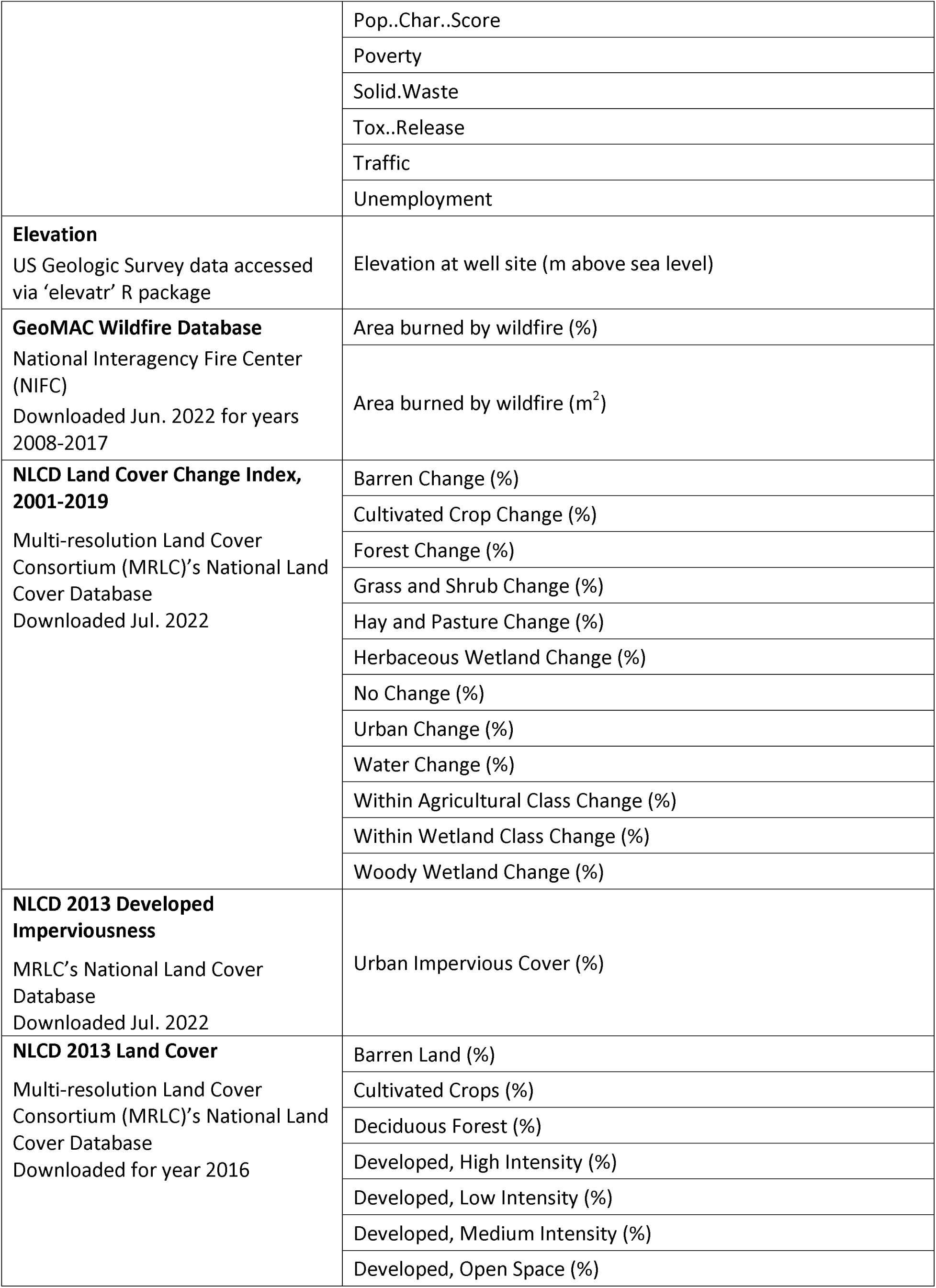
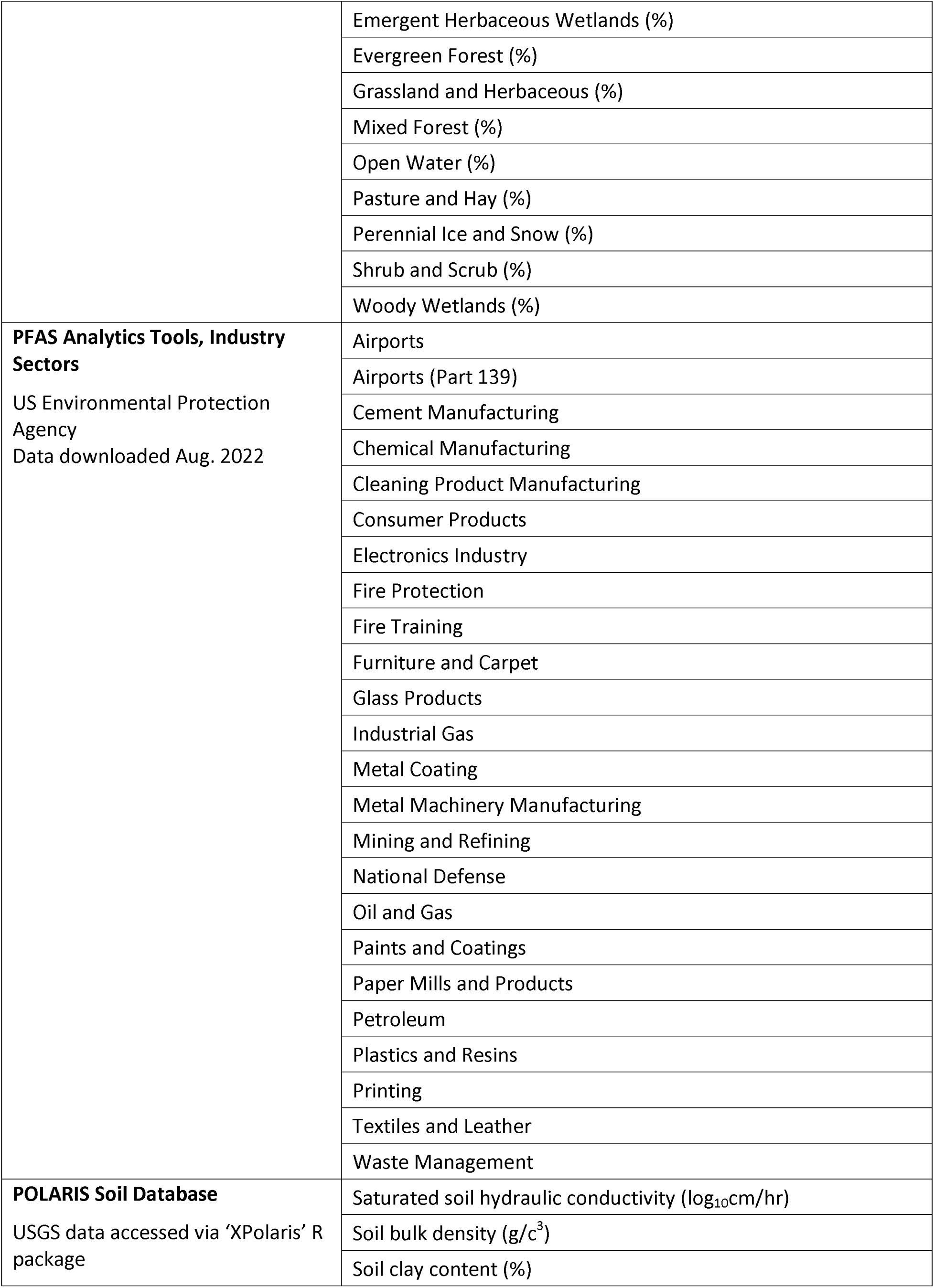
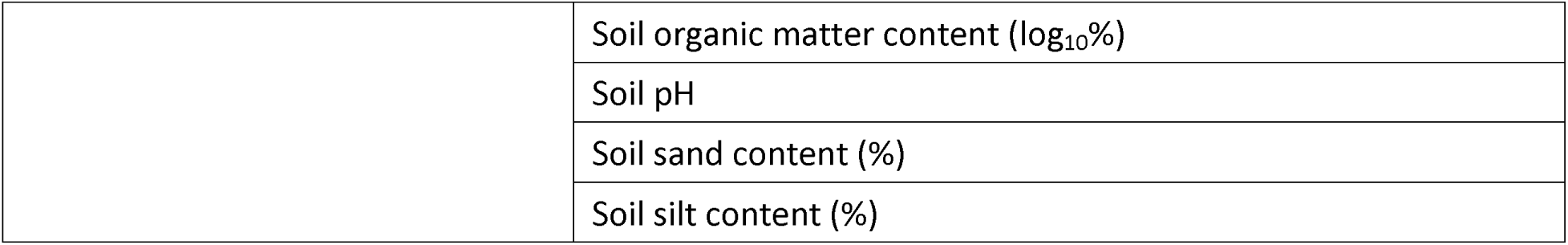
List of covariates used in the analysis.

| Data Source | Variable |
| --- | --- |
| **5-Year American Community Survey**<br>US Census Bureau data accessed via 'tidycensus' R Package<br>Downloaded for year 2015 | Age, Median |
|  | Age, Over 65 (%) |
|  | Age, Over 85 (%) |
|  | Employed in Agriculture, Forestry, Fishing, Hunting, and Mining (%) |
|  | Employed in Arts, Entertainment, Recreation, Accommodation, & Food Services (%) |
|  | Employed in Construction (%) |
|  | Employed in Educational Services, Health Care, and Social Assistance (%) |
|  | Employed in Finance, Insurance, Real Estate, Rental, and Leasing (%) |
|  | Employed in Information (%) |
|  | Employed in Manufacturing (%) |
|  | Employed in Other Services, except Public Administration (%) |
|  | Employed in Professional, Scientific, Management, Administrative, & Waste Mgmt. Services (%) |
|  | Employed in Public Administration (%) |
|  | Employed in Retail Trade (%) |
|  | Employed in Transportation, Warehousing, and Utilities (%) |
|  | Employed in Wholesale Trade (%) |
|  | Foreign Born (%) |
|  | GINI Index of Income Inequality |
|  | Health Insurance, None (%) |
|  | Health Insurance, Private (%) |
|  | Health Insurance, Public (%) |
|  | Health Insurance, Public, Medicaid (%) |
|  | Health Insurance, Public, Medicare (%) |
|  | Health Insurance, Public, VA (%) |
|  | Hispanic or Latino (%) |
|  | Households Receiving Food Stamps/SNAP (%) |
|  | Median Age of Home |
|  | Median Annual Income (USD) |
|  | Median Home Value (USD) |
|  | Median Monthly Rent (USD) |
|  | Population Living in Owner Occupied Housing (%) |
|  | Population Living in Renter Occupied Housing (%) |
|  | Population Living with a Disability (%) |
|  | Population with Bachelor's Degree or Higher (%) |
|  | Race, American Indian and Alaska Native (%) |
|  | Race, Asian (%) |
|  | Race, Black or African American (%) |
|  | Race, Native Hawaiian and Other Pacific Islander (%) |
|  | Race, Some Other Race (%) |
|  | Race, Two or More Races (%) |
|  | Race, White (%) |
|  | Sex, Female (%) |
|  | Sex, Male (%) |
|  | Total Population |
|  | Unemployment Rate (%) |
|  | Veteran (%) |
| **CalEnviroScreen, v4.0**<br>California Office of Environmental<br>Health Hazard Assessment<br>Report published Oct. 2021, data<br>downloaded Jun. 2022 | Asthma |
|  | Cardiovascular.Disease |
|  | CES.4.0.Score |
|  | Cleanup.Sites |
|  | Diesel.PM |
|  | Drinking.Water |
|  | Education |
|  | Groundwater.Threats |
|  | Haz..Waste |
|  | Housing.Burden |
|  | Imp..Water.Bodies |
|  | Lead |
|  | Linguistic.Isolation |
|  | Low.Birth.Weight |
|  | Ozone |
|  | Pesticides |
|  | PM2.5 |
|  | Pollution.Burden |
|  | Pollution.Burden.Score |
|  | Pop..Char. |
|  | Pop..Char...Score |
|  | Poverty |
|  | Solid.Waste |
|  | Tox..Release |
|  | Traffic |
|  | Unemployment |
| **Elevation**<br>US Geologic Survey data accessed via 'elevatr' R package | Elevation at well site (m above sea level) |
| **GeoMAC Wildfire Database**<br>National Interagency Fire Center (NIFC)<br>Downloaded Jun. 2022 for years 2008-2017 | Area burned by wildfire (%) |
|  | Area burned by wildfire (m<sup>2</sup>) |
| **NLCD Land Cover Change Index, 2001-2019**<br>Multi-resolution Land Cover Consortium (MRLC)'s National Land Cover Database<br>Downloaded Jul. 2022 | Barren Change (%) |
|  | Cultivated Crop Change (%) |
|  | Forest Change (%) |
|  | Grass and Shrub Change (%) |
|  | Hay and Pasture Change (%) |
|  | Herbaceous Wetland Change (%) |
|  | No Change (%) |
|  | Urban Change (%) |
|  | Water Change (%) |
|  | Within Agricultural Class Change (%) |
|  | Within Wetland Class Change (%) |
|  | Woody Wetland Change (%) |
| **NLCD 2013 Developed Imperviousness**<br>MRLC's National Land Cover Database<br>Downloaded Jul. 2022 | Urban Impervious Cover (%) |
| **NLCD 2013 Land Cover**<br>Multi-resolution Land Cover Consortium (MRLC)'s National Land Cover Database<br>Downloaded for year 2016 | Barren Land (%) |
|  | Cultivated Crops (%) |
|  | Deciduous Forest (%) |
|  | Developed, High Intensity (%) |
|  | Developed, Low Intensity (%) |
|  | Developed, Medium Intensity (%) |
|  | Developed, Open Space (%) |
|  | Emergent Herbaceous Wetlands (%) |
|  | Evergreen Forest (%) |
|  | Grassland and Herbaceous (%) |
|  | Mixed Forest (%) |
|  | Open Water (%) |
|  | Pasture and Hay (%) |
|  | Perennial Ice and Snow (%) |
|  | Shrub and Scrub (%) |
|  | Woody Wetlands (%) |
| **PFAS Analytics Tools, Industry Sectors**<br>US Environmental Protection Agency<br>Data downloaded Aug. 2022 | Airports |
|  | Airports (Part 139) |
|  | Cement Manufacturing |
|  | Chemical Manufacturing |
|  | Cleaning Product Manufacturing |
|  | Consumer Products |
|  | Electronics Industry |
|  | Fire Protection |
|  | Fire Training |
|  | Furniture and Carpet |
|  | Glass Products |
|  | Industrial Gas |
|  | Metal Coating |
|  | Metal Machinery Manufacturing |
|  | Mining and Refining |
|  | National Defense |
|  | Oil and Gas |
|  | Paints and Coatings |
|  | Paper Mills and Products |
|  | Petroleum |
|  | Plastics and Resins |
|  | Printing |
|  | Textiles and Leather |
|  | Waste Management |
| **POLARIS Soil Database**<br>USGS data accessed via 'XPolaris' R package | Saturated soil hydraulic conductivity ($\log_{10}\text{cm/hr}$) |
| | Soil bulk density ($\text{g/c}^3$) |
|  | Soil clay content (%) |
| | Soil organic matter content ($\log_{10}\%$) |
|  | Soil pH |
|  | Soil sand content (%) |
|  | Soil silt content (%) |

**Table S2.**
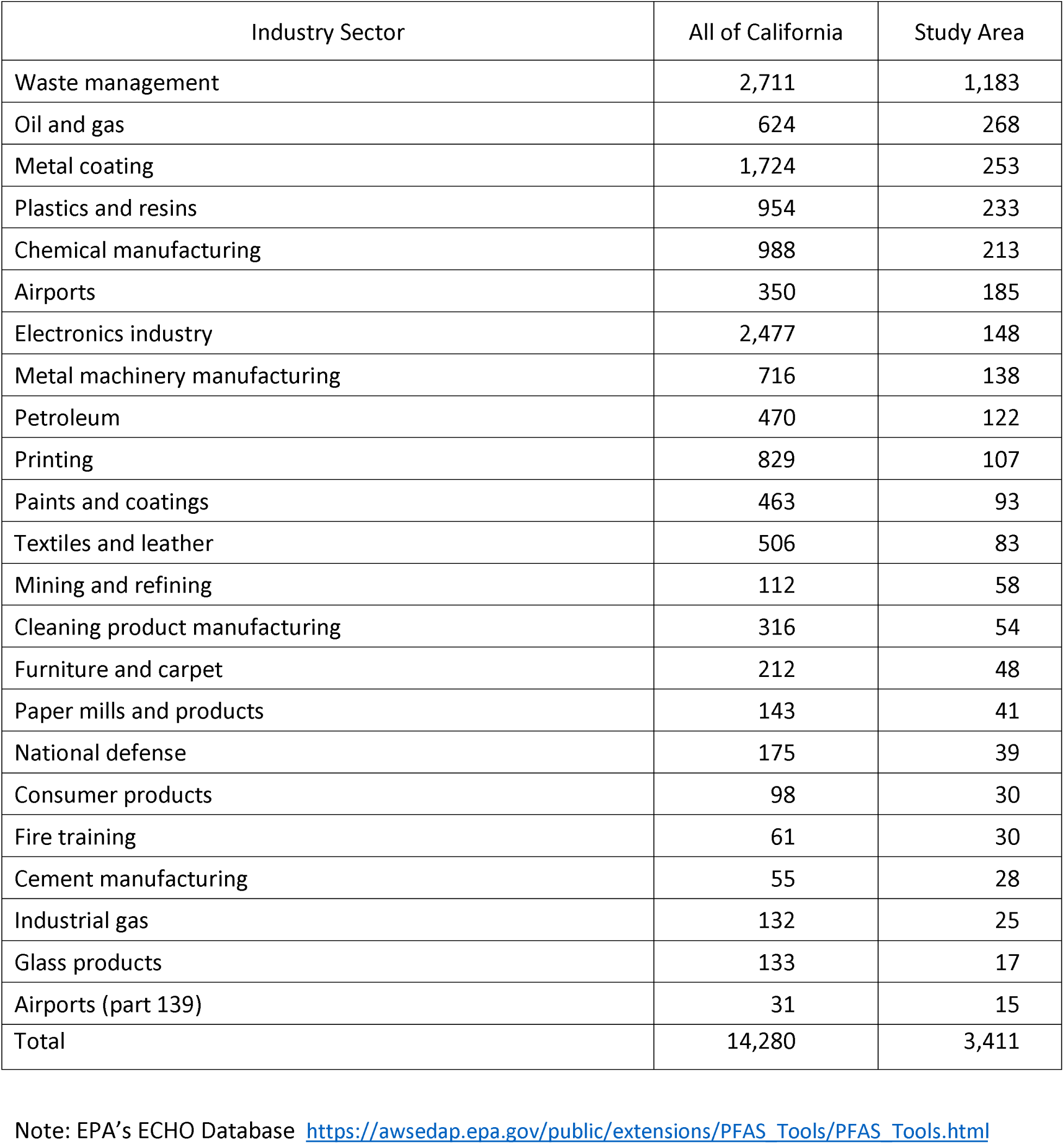
Number of PFAS Sources by 23 industrial sectors: California.

| Industry Sector | All of California | Study Area |
| --- | --- | --- |
| Waste management | 2,711 | 1,183 |
| Oil and gas | 624 | 268 |
| Metal coating | 1,724 | 253 |
| Plastics and resins | 954 | 233 |
| Chemical manufacturing | 988 | 213 |
| Airports | 350 | 185 |
| Electronics industry | 2,477 | 148 |
| Metal machinery manufacturing | 716 | 138 |
| Petroleum | 470 | 122 |
| Printing | 829 | 107 |
| Paints and coatings | 463 | 93 |
| Textiles and leather | 506 | 83 |
| Mining and refining | 112 | 58 |
| Cleaning product manufacturing | 316 | 54 |
| Furniture and carpet | 212 | 48 |
| Paper mills and products | 143 | 41 |
| National defense | 175 | 39 |
| Consumer products | 98 | 30 |
| Fire training | 61 | 30 |
| Cement manufacturing | 55 | 28 |
| Industrial gas | 132 | 25 |
| Glass products | 133 | 17 |
| Airports (part 139) | 31 | 15 |
| Total | 14,280 | 3,411 |

Note: EPA's ECHO Database https://awsedap.epa.gov/public/extensions/PFAS_Tools/PFAS_Tools.html

**Table S3.** Ten ZCTA-level covariates found to be indicative of PFAS concentrations in groundwater.

|  |
| --- |
| Covariates |
| Population percent born outside of the US |
| 2015 Population |
| Population percent living in renter-occupied unit |
| Population percent living in owner-occupied unit |
| Population percent working in manufacturing |
| Percent of area with new water or wetland landcover |
| Population percent of "Other" race |
| Population percent of American Indian or Alaskan native |
| Percent of area with new urban landcover |
| Average home age in years |

**Table S4.** Summary sampling results for 13 per- and polyfluoroalkyl substances (PFAS) species detected.

| Chemical |  | Mean and<br>(Maximum)<br>in ng/L | Number of<br>Wells with<br>PFAS<br>Detected<br>(Number<br>Wells of<br>Tested) | Number of<br>Well<br>Maximums<sup>a</sup> |
| --- | --- | --- | --- | --- |
| PFOS | (perfluorooctanoic sulfonate) | 11.1 (510.0) | 448 (1,646) | 269 |
| PFHXS | (perfluorohexanesulfonic acid) | 7.3 (690.0) | 447 (1,618) | 154 |
| PFOA | (perfluorooctanoic acid) | 6.6 (350.0) | 407 (1,647) | 67 |
| PFHA | (perfluorohexanoic acid) | 5.3 (510.0) | 359 (1,628) | 53 |
| PFBSA | (perfluorobutanesulfonic acid) | 3.2 (190.0) | 331 (1,618) | 36 |
| ADONA | (4,8-dioxa-3H-perfluorononanoic acid) | <0.1 (16.0) | 12 (1,257) | 8 |
| PFPA | (perfluoropentanoic acid) | 0.1 (6.8) | 12 (247) | 5 |
| 6:2FTS | (6:2 fluorotelomer sulfonic acid) | <0.1 (5.9) | 3 (247) | 3 |
| PFHPA | (perfluoroheptanoic acid) | 2.2 (170.0) | 241 (1,636) | 2 |
| 8:2FTS | (8:2 fluorotelomer sulfonic acid) | <0.1 (2.6) | 1 (247) | 1 |
| NETFOSAA | (N-ethyl perfluorooctane sulfonamidoacetic acid) | <0.1 (6.0) | 4 (1,610) | 1 |
| PFNA | (perfluorononanoic acid) | 0.5 (180.0) | 103 (1,636) | 1 |
| PFNDCA | (perfluorodecanoic acid) | 0.1 (17.0) | 42 (1,627) | 1 |

in groundwater wells within the study area: California, 2018-2021.
<sup>a</sup> Number of wells where this PFAS species had the highest concentration among all sampled species and thus contributed to the exposure calculations at those wells.
Note: the sum of this column (n=601) exceeds the total number of wells in study area (n=589) because 12 wells had multiple PFAS compounds tied for the maximum concentration at those wells.
Note: Four additional PFAS species (PFBTA, PFPES, PFUNDCA, NMEFOSAA) were detected in some wells, but were never the maximum PFAS species sampled at any well within the study area. Testing occurred for an additional 13 PFAS species (11CIPF3OUDS, 4:2FTS, 9CIPF3ONS, ETFOSA, HFPA-DA, MEFOSA, PFDOA, PFDSA, PFHPSA, PFNS, PFOSA, PFTEDA, PFTRIDA), but all samples in study area were below the minimum detection limit.

**Table S5.**
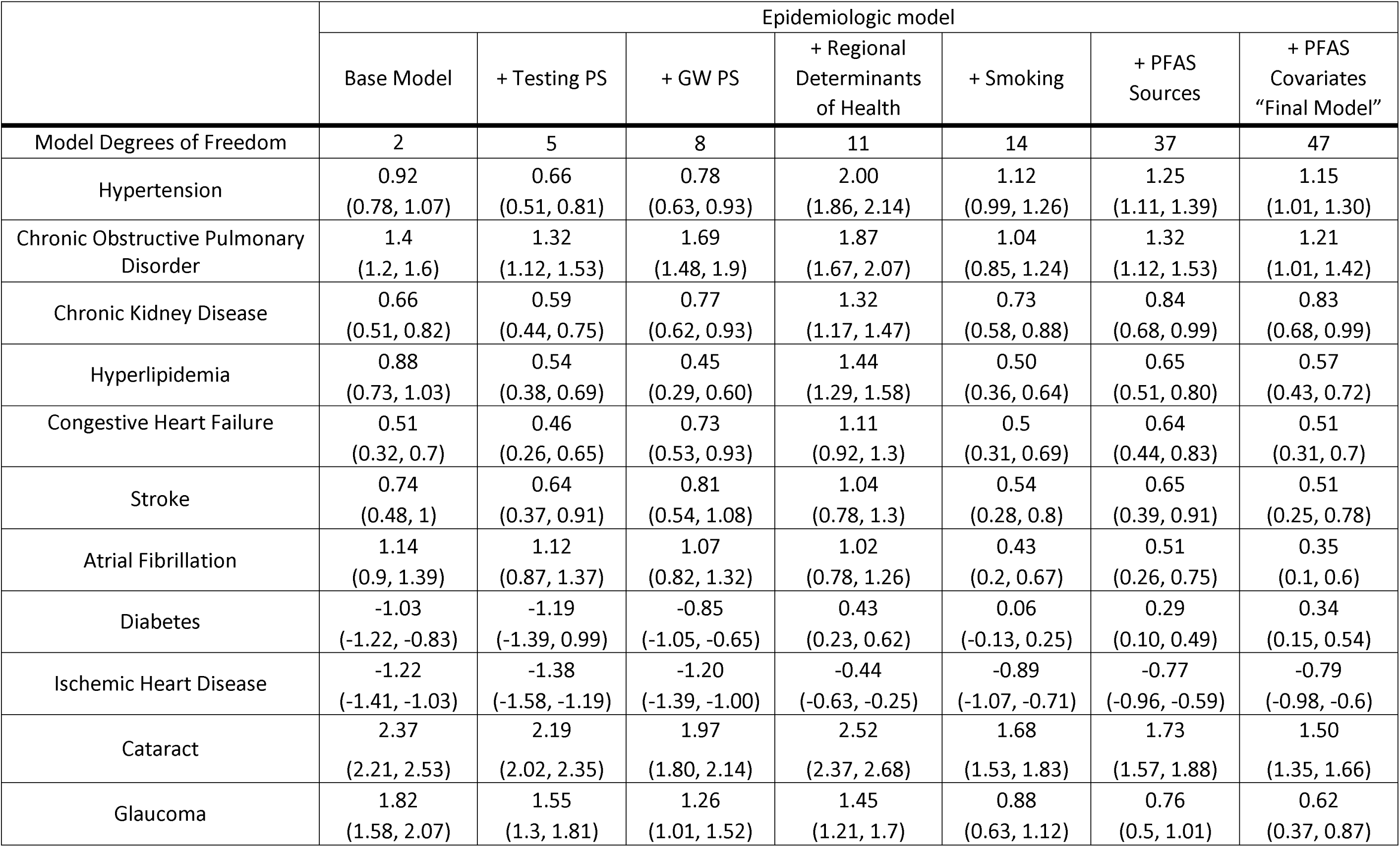
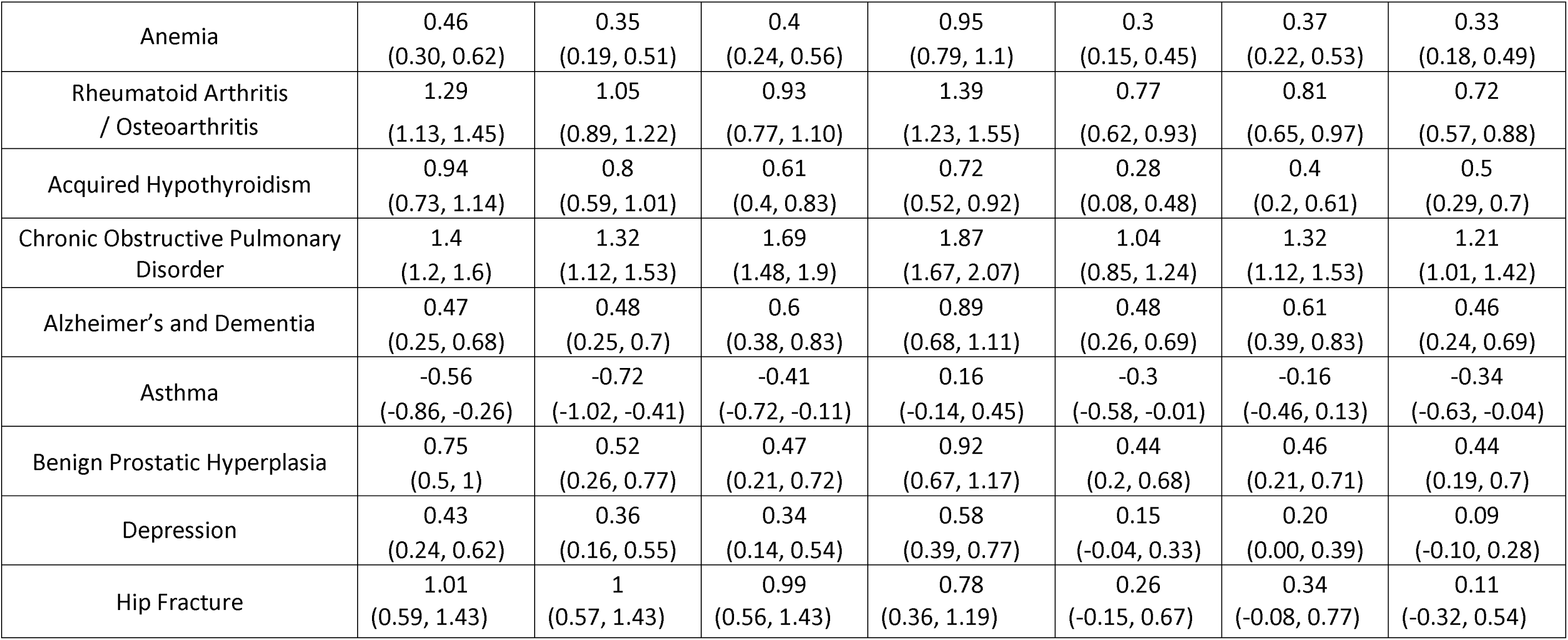
Estimated percent difference (and 95% confidence intervals) in annual incidence of 17 chronic health conditions for a 10 ng/L increment in distance weighted mean per- and polyfluoroalkyl substances (PFAS) groundwater concentration by epidemiologic model: 1,696,247 Medicare beneficiaries over age 65 living in California counties with more than 25% of drinking water from groundwater, 2011-2017.

**Figure S1.**
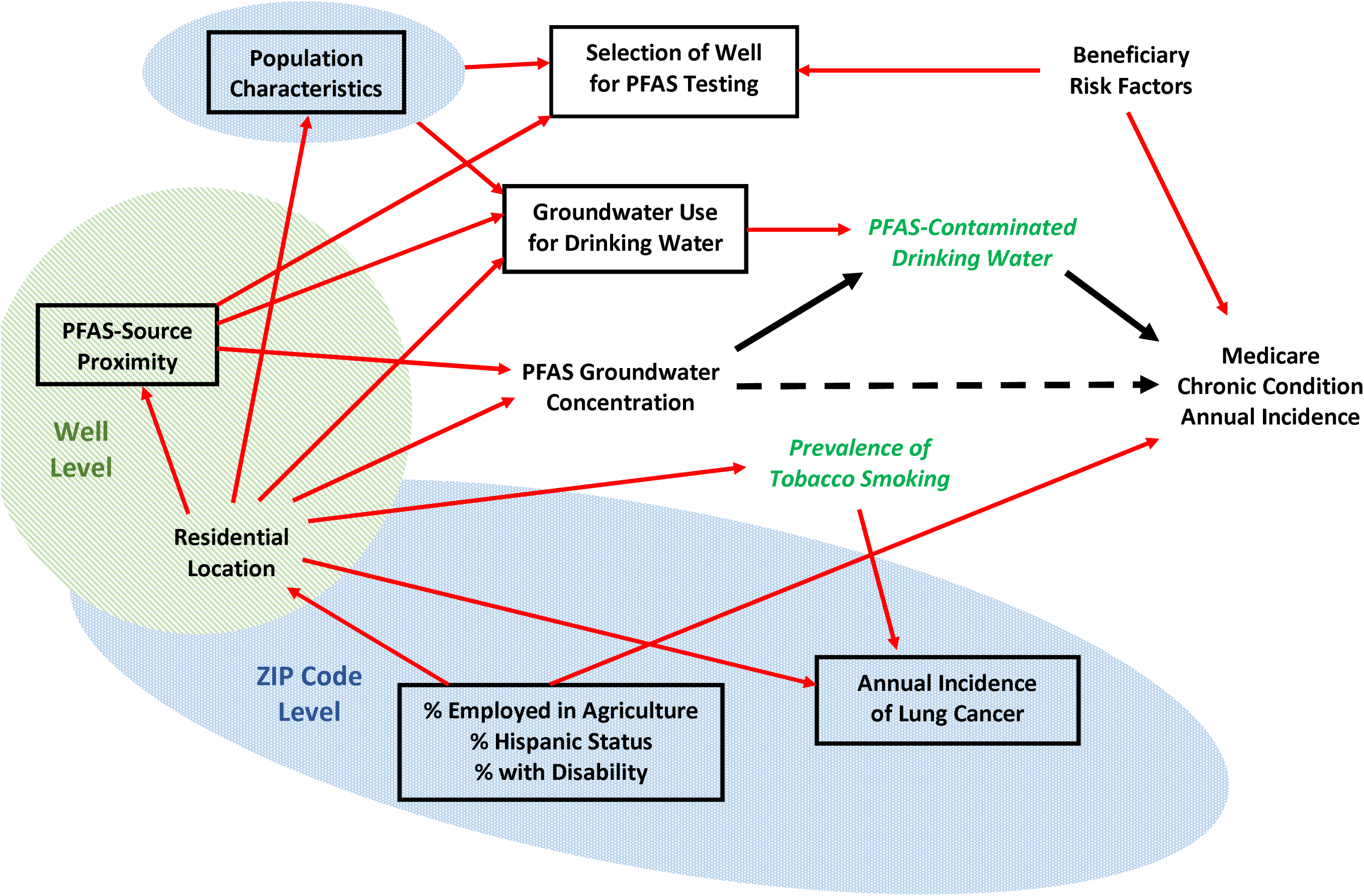
Directed Acyclic Graph (DAG) used for development of epidemiologic models.

**Figure S2.**
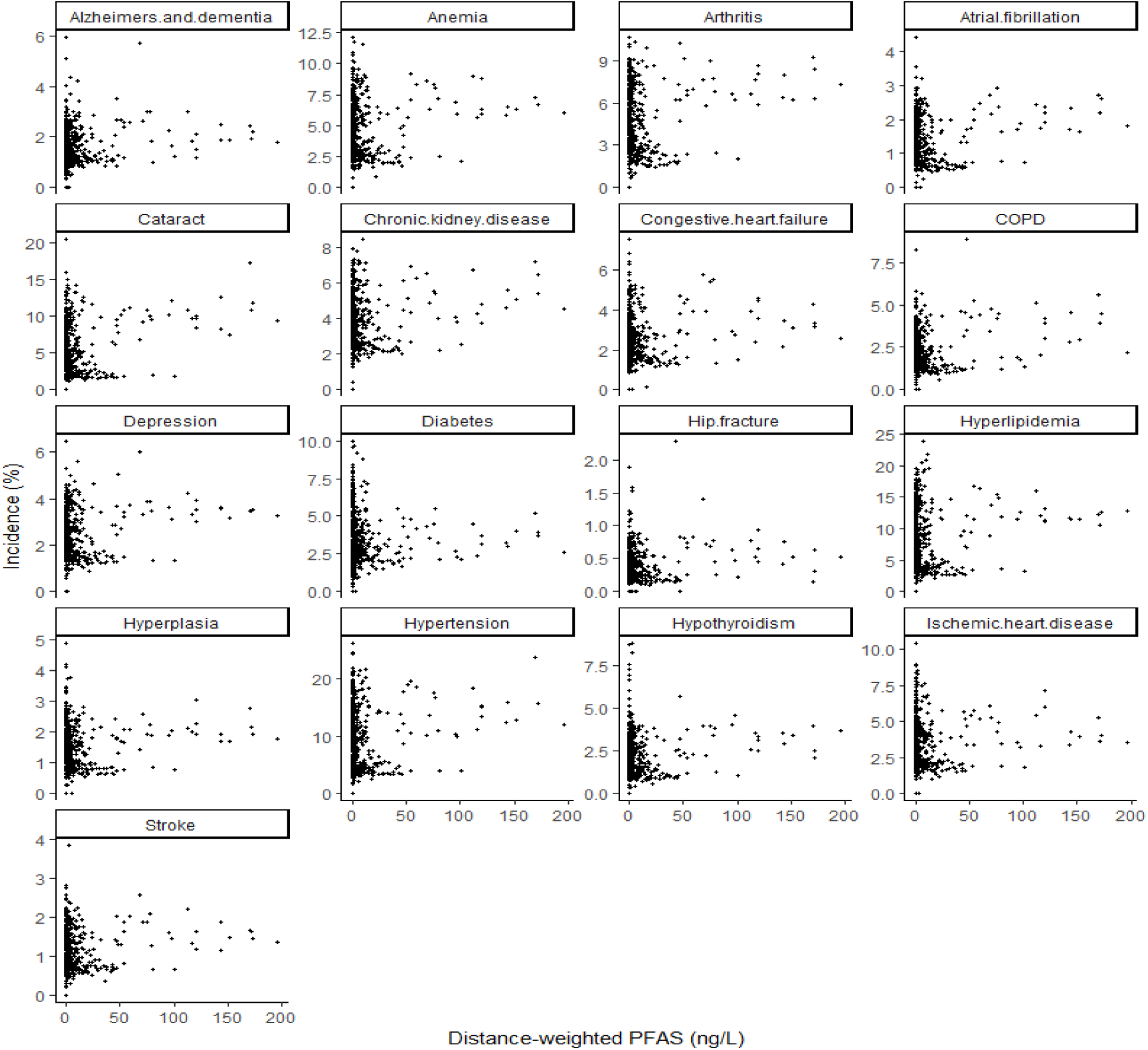
Distance-weighted mean of the well-specific maximum per- and polyfluoroalkyl substances (PFAS) concentrations (ng/L) versus 17 chronic condition incidence rates (%) by study area ZCTAs (n=523): 1,696,247 Medicare beneficiaries over age 65 living in California counties with more than 25% of drinking water from groundwater, 2011-2017.

**Figure S3.**
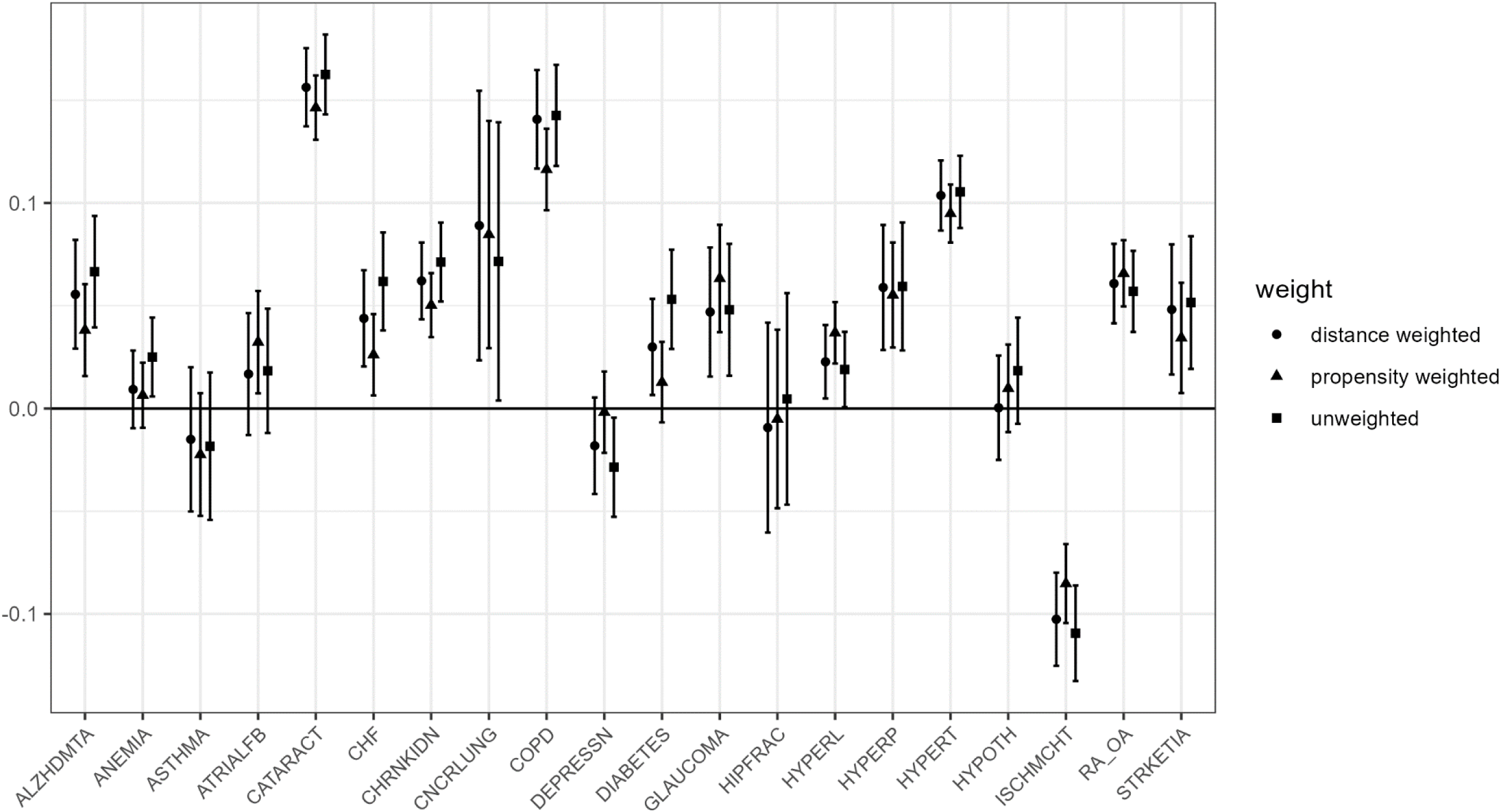
Sensitivity analysis results. Final model results for four chronic conditions split by different per- and polyfluoroalkyl substances (PFAS) exposure metrics – distance-weighted average of well maximums, distance- and propensity-weighted average of well maximums, and the unweighted average of well maximums across ZCTAs in study area without the ZCTA with a single largest PFAS value (n=522): 1,696,247 Medicare beneficiaries over age 65 living in California counties with more than 25% of drinking water from groundwater, 2011-2017.

## References

1. Avanasi, R., H. M. Shin, V. M. Vieira, D. A. Savitz and S. M. Bartell (2016). “Impact of Exposure Uncertainty on the Association between Perfluorooctanoate and Preeclampsia in the C8 Health Project Population.” Environ Health Perspect 124(1): 126–132.

2. Averina, M., J. Brox, S. Huber and A. S. Furberg (2021). “Exposure to perfluoroalkyl substances (PFAS) and dyslipidemia, hypertension and obesity in adolescents. The Fit Futures study.” Environ Res 195: 110740.

3. Birukov, A., L. B. Andersen, M. S. Andersen, J. H. Nielsen, F. Nielsen, H. B. Kyhl, J. S. Jorgensen, P. Grandjean, R. Dechend and T. K. Jensen (2021). “Exposure to perfluoroalkyl substances and blood pressure in pregnancy among 1436 women from the Odense Child Cohort.” Environ Int 151: 106442.

4. Borghese, M. M., M. Walker, M. E. Helewa, W. D. Fraser and T. E. Arbuckle (2020). “Association of perfluoroalkyl substances with gestational hypertension and preeclampsia in the MIREC study.” Environ Int 141: 105789.

5. California Office of Environmental Health Hazard Assessment. (2023). “CalEnviroScreen.” from https://oehha.ca.gov/calenviroscreen.

6. California State Water Resources Control Board. (2019). “Drinking water - public water system information. California Open Data.” Retrieved August 9, 2019, 2019, from https://data.ca.gov/dataset/drinking-water-public-water-system-information.

7. California Water Boards, S. W. R. C. B. (2023, 5/20/2023). “GAMA DATA DOWNLOADS.” from https://gamagroundwater.waterboards.ca.gov/gama/datadownload.

8. Carlson, L. M., M. Angrish, A. V. Shirke, E. G. Radke, B. Schulz, A. Kraft, R. Judson, G. Patlewicz, R. Blain, C. Lin, N. Vetter, C. Lemeris, P. Hartman, H. Hubbard, X. Arzuaga, A. Davis, L. V. Dishaw, I. L. Druwe, H. Hollinger, R. Jones, J. P. Kaiser, L. Lizarraga, P. D. Noyes, M. Taylor, A. J. Shapiro, A. J. Williams and K. A. Thayer (2022). “Systematic Evidence Map for Over One Hundred and Fifty Per- and Polyfluoroalkyl Substances (PFAS).” Environ Health Perspect 130(5): 56001.

9. Chaney, N. W., E. F. Wood, A. B. McBratney, J. W. Hempel, T. W. Nauman, C. W. Brungard and N. P. Odgers (2016). “POLARIS: A 30-meter probabilistic soil series map of the contiguous United States.” Geoderma 274: 54–67.

10. Chen, T. and C. Guestrin (2016). “XGBoost:A scalable Tree Boosting System.” Knowledge Discovery in Databases: 785–794.

11. Dewitz, J. A. and U. G. Survey. (2021). “National Land Cover Database (NLCD) 2019 Products (ver. 2.0, June 2021): U.S. Geological Survey data release.”

12. Ding, N., C. A. Karvonen-Gutierrez, B. Mukherjee, A. M. Calafat, S. D. Harlow and S. K. Park (2022). “Per- and Polyfluoroalkyl Substances and Incident Hypertension in Multi-Racial/Ethnic Women: The Study of Women’s Health Across the Nation.” Hypertension 79(8): 1876–1886.

13. Gao, X., W. Ni, S. Zhu, Y. Wu, Y. Cui, J. Ma, Y. Liu, J. Qiao, Y. Ye, P. Yang, C. Liu and F. Zeng (2021). “Per- and polyfluoroalkyl substances exposure during pregnancy and adverse pregnancy and birth outcomes: A systematic review and meta-analysis.” Environ Res 201: 111632.

14. Guelfo, J. L. and D. T. Adamson (2018). “Evaluation of a national data set for insights into sources, composition, and concentrations of per- and polyfluoroalkyl substances (PFASs) in U.S. drinking water.” Environ Pollut 236: 505–513.

15. Hamrahian, S. M. and B. Falkner (2017). “Hypertension in Chronic Kidney Disease.” Adv Exp Med Biol 956: 307–325.

16. Hollister, J., T. Shah, A. Robitaille, M. Beck and M. Johnson. (2022, 5/30/2023). “elevatr: Access Elevation Data from Various APIs.” from https://github.com/jhollist/elevatr/.

17. Hu, X. C., D. Q. Andrews, A. B. Lindstrom, T. A. Bruton, L. A. Schaider, P. Grandjean, R. Lohmann, C. C. Carignan, A. Blum, S. A. Balan, C. P. Higgins and E. M. Sunderland (2016). “Detection of Poly- and Perfluoroalkyl Substances (PFASs) in U.S. Drinking Water Linked to Industrial Sites, Military Fire Training Areas, and Wastewater Treatment Plants.” Environ Sci Technol Lett 3(10): 344–350.

18. Johnson, T. D., K. Belitz and M. A. Lombard (2019). “Estimating domestic well locations and populations served in the contiguous US for years 2000 and 2010.” Science of the Total Environment 687: 1261–1273.

19. Kim, S. H., J. H. Park, J. K. Lee, E. Y. Heo, D. K. Kim and H. S. Chung (2017). “Chronic obstructive pulmonary disease is independently associated with hypertension in men: A survey design analysis using nationwide survey data.” Medicine (Baltimore) 96(19): e6826.

20. Ma, S., C. Xu, J. Ma, Z. Wang, Y. Zhang, Y. Shu and X. Mo (2019). “Association between perfluoroalkyl substance concentrations and blood pressure in adolescents.” Environ Pollut 254(Pt A): 112971.

21. MacKinnon, D. P., J. L. Krull and C. M. Lockwood (2000). “Equivalence of the mediation, confounding and suppression effect.” Prev Sci 1(4): 173–181.

22. Meneguzzi, A., C. Fava, M. Castelli and P. Minuz (2021). “Exposure to Perfluoroalkyl Chemicals and Cardiovascular Disease: Experimental and Epidemiological Evidence.” Front Endocrinol (Lausanne) 12: 706352.

23. Moro Rosso, L. H., A. F. de Borja Reis, A. A. Correndo and I. A. Ciampitti (2021). “XPolaris: an R-package to retrieve United States soil data at 30-meter resolution.” BMC Res Notes 14(1): 327.

24. National Science and Technology Council (2023). Per- and Polyfloroalkyl Substances (PFAS) Report. I. Joint Subcommittee on Environment, and Public Health,. Washington, DC, Executive Office of the President of the United States.

25. Preston, E. V., M. F. Hivert, A. F. Fleisch, A. M. Calafat, S. K. Sagiv, W. Perng, S. L. Rifas-Shiman, J. E. Chavarro, E. Oken, A. R. Zota and T. James-Todd (2022). “Early-pregnancy plasma per- and polyfluoroalkyl substance (PFAS) concentrations and hypertensive disorders of pregnancy in the Project Viva cohort.” Environ Int 165: 107335.

26. R Core Team (2022). R: A Language and Environment for Statistical Computing. Vienna, Austria, R Foundation for Statistical Computing.

27. Radke, E. G., J. M. Wright, K. Christensen, C. J. Lin, A. E. Goldstone, C. Lemeris and K. A. Thayer (2022). “Epidemiology Evidence for Health Effects of 150 per- and Polyfluoroalkyl Substances: A Systematic Evidence Map.” Environ Health Perspect 130(9): 96003.

28. Rickard, B. P., I. Rizvi and S. E. Fenton (2022). “Per- and poly-fluoroalkyl substances (PFAS) and female reproductive outcomes: PFAS elimination, endocrine-mediated effects, and disease.” Toxicology 465: 153031.

29. Rylander, L., C. H. Lindh, S. R. Hansson, K. Broberg and K. Kallen (2020). “Per- and Polyfluoroalkyl Substances in Early Pregnancy and Risk for Preeclampsia: A Case-Control Study in Southern Sweden.” Toxics 8(2).

30. Starling, A. P., S. M. Engel, D. B. Richardson, D. D. Baird, L. S. Haug, A. M. Stuebe, K. Klungsoyr, Q. Harmon, G. Becher, C. Thomsen, A. Sabaredzovic, M. Eggesbo, J. A. Hoppin, G. S. Travlos, R. E. Wilson, L. I. Trogstad, P. Magnus and M. P. Longnecker (2014). “Perfluoroalkyl substances during pregnancy and validated preeclampsia among nulliparous women in the Norwegian Mother and Child Cohort Study.” Am J Epidemiol 179(7): 824–833.

31. Stein, C. R., D. A. Savitz and M. Dougan (2009). “Serum levels of perfluorooctanoic acid and perfluorooctane sulfonate and pregnancy outcome.” Am J Epidemiol 170(7): 837–846.

32. Tian, Y., Q. Zhou, L. Zhang, W. Li, S. Yin, F. Li and C. Xu (2023). “In utero exposure to per-/polyfluoroalkyl substances (PFASs): Preeclampsia in pregnancy and low birth weight for neonates.” Chemosphere 313: 137490.

33. US Bureau of the Census. (2023). “American Community Survey 5-Year Data (2009-2021).” from https://www.census.gov/data/developers/data-sets/acs-5year.html.

34. US Bureau of the Census. (2023). “TIGER/Line Shapefiles.” 2023, from https://www.census.gov/geographies/mapping-files/time-series/geo/tiger-line-file.html.

35. US Department of Health and Human Services. (2023). “Chronic Conditions Data Warehouse.” from https://www2.ccwdata.org/web/guest/condition-categories-chronic/.

36. US Department of Housing and Urban Development. (2023). “Geospatial Data Storefront.” from https://hudgis-hud.opendata.arcgis.com/.

37. US Environmental Protection Agency (2021). Announcement of Final Regulatory Determinations for Contaminants on the Fourth Drinking Water Contaminant Candidate List. O. o. Water. Federal Register. 86: 12272-12291.

38. US Environmental Protection Agency. (2023). “PFAS Analytic Tools.” from https://awsedap.epa.gov/public/extensions/PFAS_Tools/PFAS_Tools.html.

39. US Geological Survey. (2023). “Groundwater Ambient Monitoring & Assessment (GAMA).” from https://www.usgs.gov/centers/california-water-science-center/science/groundwater-ambient-monitoring-assessment-gama.

40. Ward-Caviness, C. K., J. Moyer, A. Weaver, R. Devlin and D. Diaz-Sanchez (2022). “Associations between PFAS occurrence and multimorbidity as observed in an electronic health record cohort.” Environ Epidemiol 6(4): e217.

41. Wen, X., M. Wang, X. Xu and T. Li (2022). “Exposure to Per- and Polyfluoroalkyl Substances and Mortality in U.S. Adults: A Population-Based Cohort Study.” Environ Health Perspect 130(6): 67007.

42. Wickham, J., S. V. Stehman, D. G. Sorenson, L. Gass and J. A. Dewitz (2021). “Thematic accuracy assessment of the NLCD 2016 land cover for the conterminous United States.” Remote Sensing of Environment 257.

43. Yu, X., D. Lyu, X. Dong, J. He and K. Yao (2014). “Hypertension and risk of cataract: a meta-analysis.” PLoS One 9(12): e114012.

